# Spatial Profiling of Patient-Matched HER2 Positive Gastric Cancer Reveals Resistance Mechanisms to Trastuzumab and Trastuzumab Deruxtecan

**DOI:** 10.1101/2024.10.29.24316248

**Authors:** Taotao Sheng, Raghav Sundar, Supriya Srivastava, Xuewen Ong, Su Ting Tay, Haoran Ma, Tomoyuki Uchihara, Benedict Shi Xiang Lian, Takeshi Hagihara, Chang Xu, Shamaine Wei Ting Ho, Kie Kyon Huang, Angie Lay Keng Tan, Michelle Shu Wen NG, Ng Shi Ya Clara, Vincenzo Nasca, Chiara Carlotta Pircher, Giovanni Randon, Silvia Giordano, Simona Corso, Jeffrey Huey Yew Lum, Ming Teh, Jimmy Bok Yan So, Jessica Gasparello, Matteo Fassan, Filippo Pietrantonio, Patrick Tan

## Abstract

**PURPOSE:** HER2-positive gastric cancer (HER2+ GC) exhibits significant intra-tumoral heterogeneity and frequent development of resistance to HER2-targeted therapies. This study aimed to characterize the spatial tumor microenvironment (TME) in HER2+ GC and identify mechanisms of resistance to HER2 blockade including trastuzumab and trastuzumab deruxtecan (T-DXd), with the goal of informing novel therapeutic strategies.

**PATIENTS AND METHODS:** We performed spatial transcriptomics on pre-and post-treatment samples from patients with HER2+ metastatic GC who received trastuzumab-based therapy. We also established patient-derived organoids (PDOs) to investigate mechanisms of trastuzumab resistance *in vitro*.

**RESULTS:** *ERBB2*-high tumor regions were found to be "immune cold", characterized by low *PD-L1* expression and reduced lymphocyte infiltration. We identified two distinct mechanisms of acquired trastuzumab resistance: epithelial-mesenchymal transition (EMT) and upregulation of the endoplasmic reticulum-associated protein degradation (ERAD) pathway. EMT-positive tumors showed increased expression of immune checkpoints, including *PD-L1*, and the chemokine *CCL2*. Non-EMT tumors exhibited upregulation of the ERAD pathway, highlighting it as a potential therapeutic target. Importantly, we observed increased expression of the promising therapeutic target CLDN18.2, in trastuzumab-resistant tumors. Additionally, loss of HLA was identified as a potential mechanism of resistance to trastuzumab deruxtecan (T-DXd).

**CONCLUSION:** Our spatial profiling study reveals distinct TME features and resistance mechanisms in HER2+ GC, providing a valuable resource for future research and therapeutic development. The identification of potential therapeutic targets, such as CLDN18.2, may pave the way for novel treatment strategies to overcome resistance and improve outcomes for patients with HER2+ GC.

## Introduction

Gastric cancer (GC) is the fifth most common malignancy globally contributing significantly to worldwide cancer-related mortality^1^. Approximately 15–20% of GCs exhibit overexpression and/or amplification of human epidermal growth factor receptor 2 (HER2; also known as *ERBB2)*, termed HER2-positive GC (HER2+ GC)^2^. Although cancer cell-intrinsic HER2 signaling in driving growth and proliferation has been well-studied, the impact of HER2 signaling on the tumor immune microenvironment remains unclear. Some studies suggest that HER2+ GCs have lower immune cell infiltration and expression of immune checkpoints including *PD-L1*^3,4^, while other groups have reported no association between HER2+ GC status and tumor-infiltrating CD8^+^ T cells or *PD-L1* expression^5^. Some of these disparities may be due to intra-tumoral HER2 heterogeneity, observed in over half of HER2+ GC patients^6–8^.

HER2 has been successfully exploited for targeted therapy using monoclonal antibodies such as trastuzumab^9^. Unfortunately, for many trastuzumab-treated patients, despite initial responses disease progression occurs in most cases within one year due to acquired resistance^9,10^. Trastuzumab deruxtecan (T-DXd) is a promising second-or third-line treatment following trastuzumab therapy^11,12^. However, most patients eventually also develop T-DXd acquired resistance. There is an urgent need to uncover and overcome HER2 resistance mechanisms to improve advanced GC patient outcomes.

Previous research has identified several potential mechanisms of trastuzumab resistance in HER2+ GC, including secondary mutations of HER2, co-amplifications of RTKs, reactivation of downstream pathways, and epigenetic changes^13–19^. However, these mechanisms have primarily been studied *in vitro* and lack validation in clinical samples^20^, with most research focusing on bulk tumor analyses. In this study we utilized the NanoString Digital Spatial Profiler (GeoMx DSP) technology^21^ to analyze 46 samples from 30 GC patients spanning different stages of anti-HER2 treatment. We find that *ERBB2*-high tumor regions are immune-cold both within and across patients. Additionally, we delineated two separate mechanisms of acquired resistance to trastuzumab in HER2+GC, each associated with distinct molecular pathways. This extensive spatial dataset provides a valuable resource for future HER2+ GC research.

## Methods

### Patient sample collection

FFPE tissue samples were obtained from patients (PT1-15) diagnosed with HER2+ metastatic gastric cancer (GC) who received anti-HER2 standard therapies at the Fondazione IRCCS Istituto Nazionale dei Tumori, Milan, Italy, and patients (PNT1-15) diagnosed with gastric adenocarcinoma undergoing surgical resection or endoscopy at the National University Hospital, Singapore. This study was conducted in accordance with the Declaration of Helsinki. Written informed consent was obtained from all study subjects prior to enrolment into the study. Further details on patient information and clinical data collection can be found in the **Supplementary Methods**.

### Spatial profiling

Spatial transcriptomics was performed on FFPE tissue samples using the NanoString GeoMx DSP platform. Spatial proteomics was performed using the COMET platform (Lunaphore Technologies SA) with a panel of antibodies targeting stromal and immune markers. Image analysis and cell segmentation were performed using QuPath and cellXpress software. Details of the spatial profiling methods are available in the **Supplementary Methods**.

### Establishment of Organoids and Identification of HER2+ Organoids

Patients at the National University Hospital, Singapore were enrolled in this study following approval from the local ethics board. Specimens were obtained via endoscopic biopsy. Organoids were established using a protocol adapted from Nanki et al.^22^ with some modifications. Immunofluorescence (IF) staining was performed to evaluate the expression of HER2 in HER2+ organoids. Details available in **Supplementary Methods.**

### Statistical Analysis

Statistical analyses were conducted using the R software (version 4.1.2). Further details on statistical methods and data analysis are provided in the **Supplementary Methods**.

## Results

### Spatial Transcriptomics Highlights HER2+ Tumor Regions as Immune Cold

We assembled a cohort of 30 GC patients, including 15 patients (PT1-PT15) with HER2+ metastatic GC receiving trastuzumab with chemotherapy as first-line treatment (“trastuzumab”) (**Figure 1A**). Among these, 7 patients had a partial response (PR), 6 had stable disease (SD), 1 experienced disease progression, and 1 had unavailable information (**Figure 1B**). Additionally, 6 patients subsequently received T-DXd as second-line therapy. The HER2+ tumor samples were a mix of surgical resection samples, endoscopic biopsies and core biopsies from gastric or metastatic sites (**Supplementary Table 1**). We also included 15 additional patients with locally advanced GC undergoing primary tumor resections (PNT1-PNT15), who did not receive anti-HER2 based therapy, with one patient identified as HER2+ during the study (**Supplementary Table 2**). From the 30 patients, we generated spatial transcriptomic profiles (NanoString Digital Spatial Profiler; DSP) of 1,582 ROIs across 46 samples (**Figure 1A**; median 16 ROIs per sample). Each ROI (comprising ∼200-400 nuclei) was meticulously curated and annotated into tumor, stroma, immune, and adjacent normal epithelial regions by a qualified pathologist (SS). Annotations were based on cell morphology and fluorescent markers (PanCK, SMA and CD45) targeting the predominant cell type within each ROI (**Figure 1C**). Cell type deconvolution analysis confirmed high purity of ROIs, with tumor ROIs having the highest epithelial components, stroma ROIs the highest mesenchymal components, and immune ROIs the highest immune components (P<0.001, Wilcoxon test, **Supplementary Figure 1A**). Each ROI generated ∼3,600 measurable genes (Interquartile Range, IQR: 2,051 to 5,410) (see **Methods**). The 46 samples were divided into three sub-cohorts: pre-trastuzumab (PT1-PT15 and PNT1-PNT15; 29 samples), post-trastuzumab (14 samples), and post-T-DXd (3 samples, the latter two sub-cohorts drawn from PT1-PT15).

**Figure 1.**
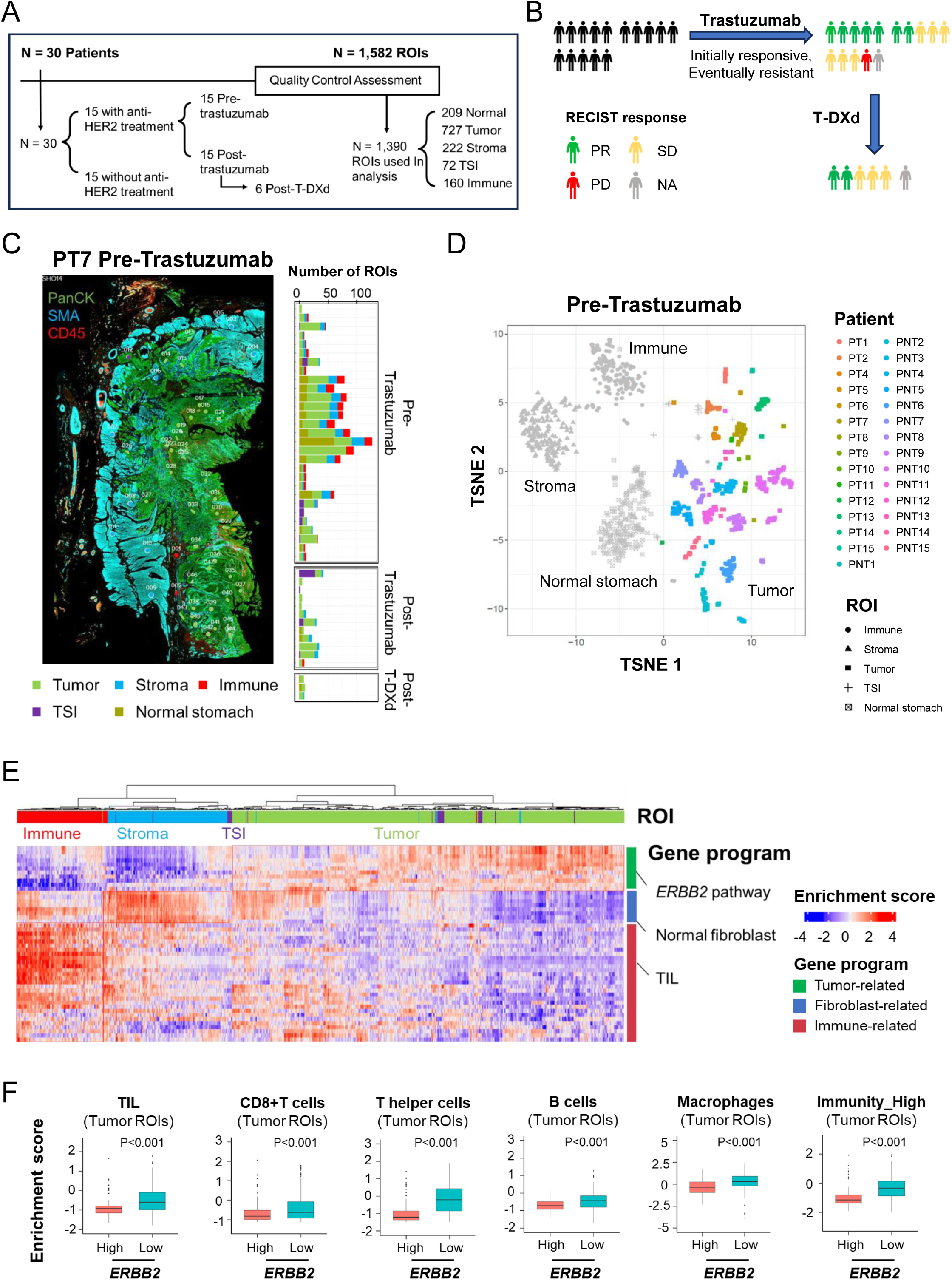
Spatially revolved tumor microenvironment (TME) in gastric cancer. A) Schematic representation of the study datasets of digital spatial profiling (DSP) from HER2+ and HER2-negative gastric cancer (GC). The GC DSP cohort comprises over 1,500 manually selected regions of interest (ROIs) from 30 GC patients, with spatial transcriptomes profiled using the NanoString GeoMx platform. B) Flow diagram depicting clinical response to HER2 blockade therapy in 15 HER2+ patients. Patients are color-coded according to their corresponding RECIST response. C) The stained DSP slide (left) from the pre-trastuzumab sample of the patient PT7. Each dot represents a ROI, with different colors denoting distinct cell types (bottom). Histogram (right) showing the number of ROIs in each patient across the pre-trastuzumab, post-trastuzumab, and post-T-DXd sub-cohorts. D) t-SNE plots of ROIs from pre-trastuzumab samples, distinguished by cell type and patient origin. Tumor ROIs are color-coded according to individual patients, whereas non-tumor ROIs are shown in grey. Each point on the t-SNE plot represents a single ROI. E) Heatmap displaying ssGSEA-based enrichment scores of various gene programs in ROIs from pre-trastuzumab samples. The immune-related gene program TIL refers to tumor infiltrating lymphocytes. F) Comparison of ssGSEA enrichment scores for various immune-related gene programs between tumor ROIs from *ERBB2*-high and *ERBB2*-low GCs. P values were calculated using the Wilcoxon test.

First, we focused on the pre-trastuzumab sub-cohort of HER2+ GC and compared them against HER2-negative GCs (1200 ROIs). UMAP analysis showed patient-specific clustering for tumor ROIs, while non-tumor ROIs clustered by category (eg stroma, immune) (**Figure 1D**). This pattern is consistent with previous findings where tumor expression profiles are frequently patient-specific patterns due to individualized tumor aneuploidy^23,24^. HER2+ tumors exhibited significantly higher levels of *ERBB2* expression and *ERBB2* pathway enrichment in tumor ROIs compared to HER2-negative tumors (P<0.001, Wilcoxon test) (**Figure 1E** and **Supplementary Figure 1B)**. We categorized tumors into *ERBB2*-high, -medium, and -low groups, based on average *ERBB2* transcript expression levels in tumor ROIs (**Supplementary Figure 1C**). Previous studies have reported that other RTKs (eg *EGFR*, *MET*) and cell cycle genes (eg *CDK6*, *CCNE1*) are often co-amplified in HER2+ GCs^16,17,25^. Indeed, *ERBB2*-high tumors exhibited significantly higher levels of *EGFR/CCNE1/CDK4/CDK6/CDK12* co-expression in tumor ROIs compared to *ERBB2*-low tumors (P<0.01, Wilcoxon test**, Supplementary Figure 1D**).

We studied associations between *ERBB2* expression and the TME. Tumor ROIs from *ERBB2*-high GCs exhibited significantly lower expression of tumor infiltrating lymphocytes (TIL) and lymphocyte gene signatures (such as CD8^+^ T cells, T helper cells, B cells, and Macrophages) compared to *ERBB2*-low tumors (P<0.001, Wilcoxon test, **Figure 1F**), a finding subsequently confirmed by cell deconvolution analysis (**Supplementary Figure 1E**). This inverse association was specific to tumor ROIs and not observed in stroma ROIs or immune ROIs (**Supplementary Figure 1F**). *PD-L1* expression was low in *ERBB2*-high tumor ROIs (**Supplementary Figure 1G)**. *ERBB2*-high tumor ROIs also exhibited low expression of recently published gene signatures associated with an immunity-high GC subtype (**Figure 1F**), suggesting these tumors are immune-cold.

We validated our findings using bulk transcriptome data from TCGA, ACRG, and GASCAD cohorts^26–28^. In TCGA, *ERBB2*-high tumors similarly demonstrated significantly lower expression of TIL signatures compared to *ERBB2*-low tumors (**Supplementary Figure 1H**). Using ESTIMATE, a bioinformatic method for assessing tumor immune components, *ERBB2*-high tumors exhibited significantly lower immune scores (P<0.001, Wilcoxon test) (**Supplementary Figure 1I**). These findings were also observed in ACRG and GASCAD (**Supplementary Figure 1H**). XCELL deconvolution on TCGA GCs revealed significant negative correlations between *ERBB2* expression and infiltration of CD8^+^ T cells, CD4^+^ T cells, B cells, and macrophages (**Supplementary Figure 1J)**. These spatial and bulk analyses confirm that tumor regions from *ERBB2*-high tumors are associated with an ‘immune-cold’ local TME.

### Within-tumor Variegation in HER2-positivity Recapitulates Across-Patient Heterogeneity

Histopathological studies indicate that HER2 expression in HER2+ GCs is often heterogeneous across tumor regions^6,29^. This variability has led to clinical guidelines recommending multiple biopsies to confirm HER2 status, potentially influencing trastuzumab resistance^7,8,30^. Our analysis confirmed significant *ERBB2* expression variability within the same tumor (**Supplementary Figure 2A)**. Spatial autocorrelation in two representative HER2+ GCs (PT13 and PT14) both showed that tumor ROIs with similar *ERBB2* levels were spatially clustered (PT13: Moran’s *I*=0.40, P=0.004; PT14: Moran’s *I*=0.57, P=0.009; **Figure 2A**).

**Figure 2.**
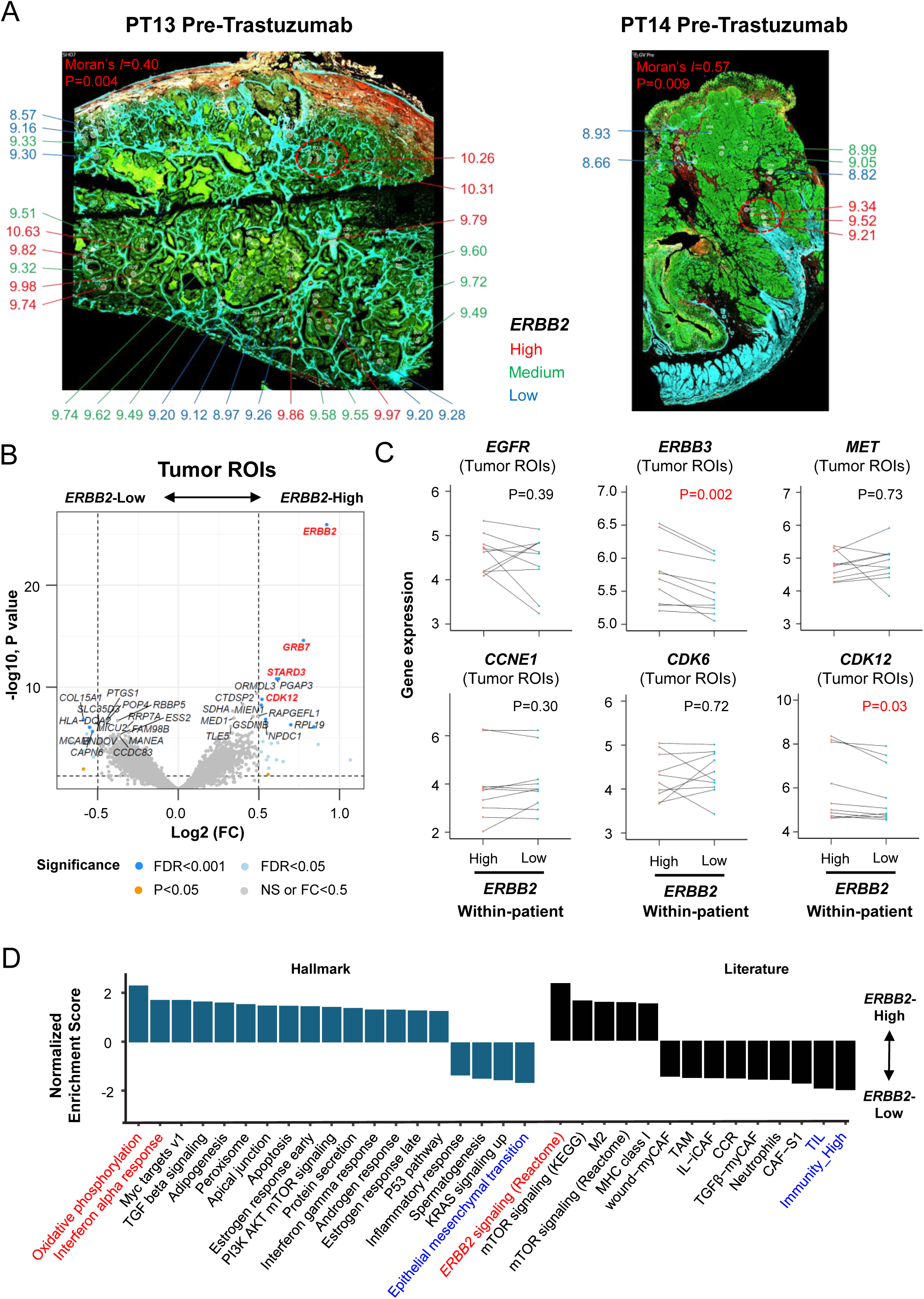
Intra-tumor HER2 heterogeneity in HER2+ gastric cancer. A) Stained DSP slides from the pre-trastuzumab sample of the patients PT13 and PT14 displaying the spatial distribution of tumor ROIs with varying *ERBB2* expression levels. *ERBB2* expression levels are color-coded according to their respective expression groups. *ERBB2*-high tumor ROIs are colored in red, *ERBB2*-medium tumor ROIs are colored in green, and *ERBB2*-low tumor ROIs are colored in blue. Moran’s indexes and P values were calculated by the Moran’s *I* test. B) Volcano plot contrasting gene expression between *ERBB2*-high and *ERBB2*-low tumor ROIs from the same patient, with axes indicating Log2 fold changes (FC) and significance levels. C) Difference in the average expression of RTKs and cell cycle genes between *ERBB2*-high and *ERBB2*-low tumor ROIs from the same patient. P values were calculated using the paired t test. D) Summary of enrichment in hallmark gene sets and literature gene signatures using differential analysis of expression data between *ERBB2*-high and *ERBB2*-low tumor ROIs from the same patient. All gene sets with P<0.05 are shown.

Comparing *ERBB2*-high and *ERBB2*-low tumor ROIs from the same patient, we identified 21 genes with significantly higher expression in *ERBB2*-high tumor ROIs (absolute log2 fold change (FC)>0.5, FDR<0.05). Reassuringly, these included *ERBB2* and other Chromosome 17q12-21 genes such as *GRB7*, *STARD3* and *CDK12* (**Figure 2B**), supporting *ERBB2* genomic amplification. *ERBB2*-high ROIs exhibited upregulation of the *ERBB2* pathway (P=0.03, paired t test) (**Supplementary Figure 2B**) and significantly higher expression of *ERBB3* (P=0.002, paired t test) (**Figure 2C**). Interestingly, *ESTY1* and *SMARCC2*, neighboring *ERBB3* on chromosome 12q13, also showed higher expression, suggesting that in some cases, the increased expression of *ERBB3* may also be contributed, at least in part, by co-amplification (**Supplementary Figure 2C**).

We curated a comprehensive resource of gene sets, including Hallmark and literature gene sets from bulk or single-cell RNA-seq^31–35^ (**Supplementary Table 3**). *ERBB2*-high tumor ROIs exhibited significantly increased expression of gene sets associated with oxidative phosphorylation, IFNα activity, and *ERBB2* signaling. Similar to the across-patient analysis (**Figure 1**), gene sets associated with EMT, TIL, and Immunity-high signatures showed reduced expression in *ERBB2*-high tumor ROIs (P<0.05, **Figure 2D**). Overall, these results demonstrate concordance in the HER2+ immune TME at both intra-patient and inter-patient resolution.

### EMT is a Major Feature of Trastuzumab Acquired Resistance

We focused on the 15 HER2+ patients receiving trastuzumab. Of these, 7 patients had a PR, 6 had SD, 1 experienced PD, and 1 had unavailable information (**Figure 3A**). The overall median duration of response to trastuzumab was 251 days (83-1,039 days) **(Supplementary Figure 3A)**. All patients eventually developed resistance. In pre-trastuzumab samples, patients with an HER2 IHC/FISH score of 3+, high *CDK12* expression, high *HLA-A* expression, low *ERBB3*, and low *CCNE1* expression were associated with a prolonged response duration (**Supplementary Figure 3B**). Some of these baseline predictors of trastuzumab response have been previously reported^9,13,36^, attesting to the fidelity of our study cohort.

**Figure 3.**
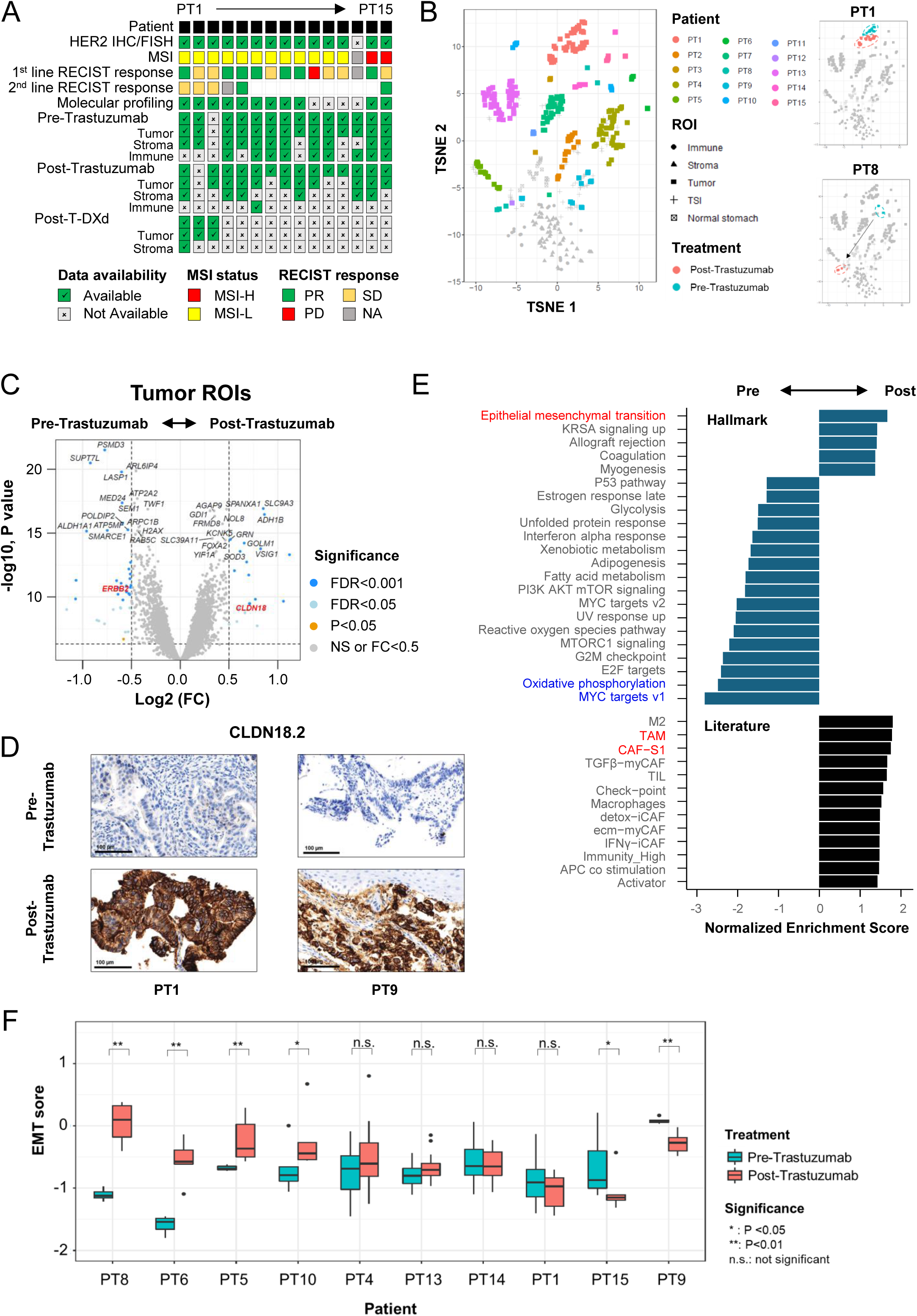
A subset of trastuzumab resistant tumors show up-regulation of EMT pathway. A) Summary of molecular diagnosis, therapy response, and DSP data in HER2+ patients. B) t-SNE plots (left) of ROIs from pre-and post-trastuzumab samples in HER2+ patients, distinguished by cell type and patient origin. Tumor ROIs are color-coded according to individual patients, whereas non-tumor ROIs are shown in grey. Each point on the t-SNE plot represents a single ROI. t-SNE plots (right) of ROIs from pre-and post-trastuzumab samples in patients PT1 and PT8, with tumor ROIs color-coded according to the treatment timeline. C) Volcano plot comparing gene expression between tumor ROIs from pre-and post-trastuzumab samples of the same patient. The axes represent Log2 FC and significance levels. D) Anti-CLDN18.2 immunohistochemistry of pre-and post-trastuzumab samples in HER2+ patients. E) Summary of enrichment in hallmark gene sets and literature gene signatures using differential analysis of expression data between tumor ROIs from pre-and post-trastuzumab samples of the same patient. All gene sets with P<0.05 are shown. F) Comparison of EMT enrichment sores between tumor ROIs from pre-and post-trastuzumab samples in each patient. Patients are ordered according to the changes in average EMT scores of tumor ROIs. P values were calculated using the Wilcoxon test.

Post-trastuzumab samples from patients allowed us to study GC molecular features after resistance. Genomic profiling of post-trastuzumab samples revealed that most frequent genetic alterations observed post-trastuzumab were *TP53*, *PIK3CA*, and *CDK12* mutations (**Supplementary Figure 3C**). Amplifications of *ERBB2*, *KRAS*, *CCNE*, *MET*, and *CDK6* were also present. These genetic alterations have been reported to be associated with trastuzumab resistance^13,37^.

To study transcriptomic alterations associated with trastuzumab resistance, we evaluated spatial transcriptomics data of 14 pre-and 14 post-trastuzumab samples (13 paired; **Figure 3A**). Notably, while tumor ROIs from some patients (eg PT1) showed high similarity between pre-and post-trastuzumab samples (**Figure 3B**, **Supplementary Figure 3D**), tumor ROIs from other patients (eg PT8), displayed a significant shift in transcriptomic profiles. *ERBB2* expression decreased significantly after trastuzumab (**Figure 3C**, **Supplementary Figure 3E**), consistent with reported HER2 losses post-trastuzumab^38–40^. Importantly, these transcriptional changes were not caused by alterations in tumor cell purity, as epithelial cell proportions remained stable post-trastuzumab for most GCs (**Supplementary Figure 3F**).

Comparing tumor ROIs from pre-and post-trastuzumab samples on a patient-wise basis, we identified 20 genes with significantly higher expression in post-trastuzumab samples (**Figure 3C**). Of potential clinical relevance, one of these genes was *CLDN18*, as CLDN18.2 has been identified as a promising GC therapeutic target^41^. We performed validation IHC analysis and confirmed a significant upregulation of CLDN18.2 following trastuzumab treatment (P=0.04, **Figure 3D**, **Supplementary Figure 3G**). Specifically, 16.7% (2/12) of pre-trastuzumab samples were CLDN18.2-positive, compared to 50% (6/12) of post-trastuzumab samples (**Supplementary Figure 3H**). Pathway analysis revealed significant increases in the expression of EMT, tumor-associated macrophage (TAM), and cancer-associated fibroblasts (CAF)-S1 signatures, while *MYC* targets and oxidative phosphorylation genes showed reduced expression (**Figure 3E**). The Hallmark EMT pathway was the top positively enriched pathway in post-trastuzumab samples, which was further confirmed using a published GC EMT gene set^42^ (P=1.49 × 10^-4^) (**Supplementary Figure 3I**). Notably, samples could be separated into two subsets, with about a third of samples (PT5, PT6, PT8 and PT10) showing significant increases in EMT-related gene expression post-treatment, while the others exhibited little changes or even decreases (**Figure 3F**). Notably, trastuzumab treatment did not significantly alter mesenchymal proportions in tumor ROIs for most patients, suggesting that the observed changes in EMT scores are unlikely attributable to alterations in mesenchymal proportions (**Supplementary Figure 3J**). This suggests that a substantial proportion of HER2+ GCs are likely to upregulate the EMT pathway post HER2 blockade.

Spatial data also enabled us to assess trastuzumab’s impact on tumor-associated stromal and immune compartments. In stroma ROIs, 84 genes, including the CAF marker *PDGFRB*, showed increased expression post-trastuzumab (**Supplementary Figure 3K**). Pathway analysis demonstrated a significant upregulation of EMT, complement, TGFβ signaling pathways, and gene signatures associated with TGFβ-myCAF, TAM, and Myeloid-derived suppressor cells (MDSCs) (**Supplementary Figure 3L**). In immune ROIs, 160 genes, including the inhibitory chemokine *CXCL3*, were upregulated post-trastuzumab (**Supplementary Figure 3M**). Pathway analysis demonstrated a significant upregulation of TNF-α/NF-κB, hypoxia, EMT signaling pathways, and gene signatures associated with inhibitory chemokines, chemokine receptor (CCR), immune checkpoints, and MDSCs (**Supplementary Figure 3N**). These results show that trastuzumab therapy can induce gene expression changes not only in tumor cells, but in the TME.

### *PDL1* and *CCL2* Upregulation in Tumors Undergoing EMT-associated Trastuzumab Resistance

In the EMT-positive group, differential expression analysis identified 1,732 genes with significantly higher expression in tumor ROIs of post-trastuzumab samples, including the mesenchymal gene markers *COL1A1* and *COL1A2* (**Supplementary Figure 4A**) and EMT-associated transcription factors (*TEAD2*, *ZEB2*; P=0.02, paired t-test, **Figure 4A**). In contrast, tumors not undergoing EMT (“non-EMT GCs”) showed no significant differences in *TEAD2* or *ZEB2* expression from pre-to post-treatment (**Figure 4A**).

**Figure 4.**
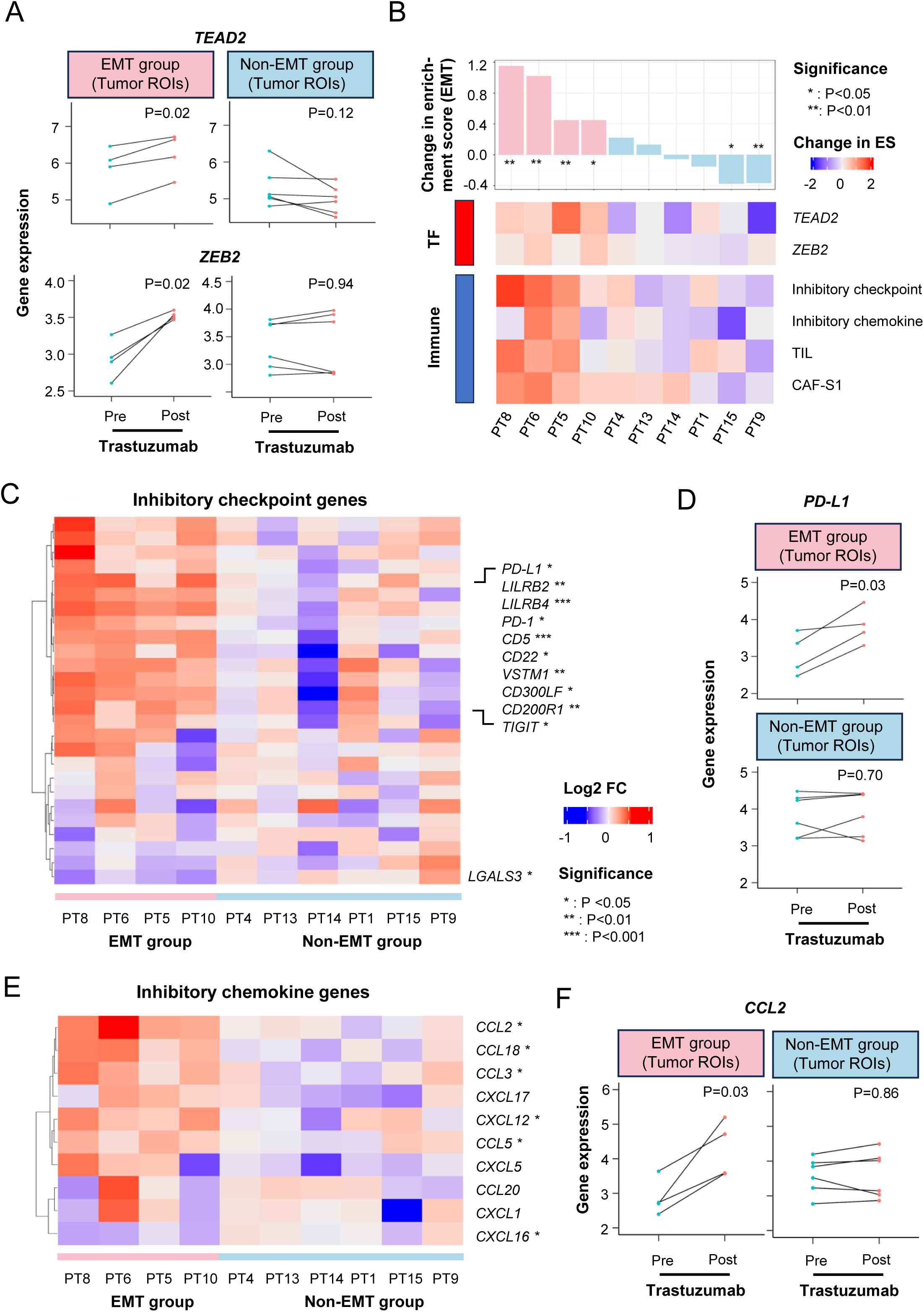
Tumors undergoing EMT-associated trastuzumab resistance have elevated expression of inhibitory immune signatures. A) Changes in the average expression of EMT-associated transcription factors *TEAD2* and *ZEB2* between tumor ROIs from pre-and post-trastuzumab samples in patients classified into the EMT and non-EMT groups. P values were calculated using the paired t test. B) Change in the average enrichment scores of key differentially regulated gene sets in tumor ROIs from ‘‘EMT’’ and ‘‘non-EMT’’ patient samples (pearson correlation P<0.05). The data are ordered according to the change in enrichment scores of the EMT signature. C) Heatmap of the log2 fold changes in the average expression of inhibitory immune checkpoints between tumor ROIs from post-and pre-trastuzumab samples. Rows represent individual immune checkpoints, and columns represent patients. Colors indicate the log2 fold change values between post-and pre-trastuzumab samples. Checkpoints that exhibited significant changes in tumors undergoing EMT post-trastuzumab were highlighted. P values were calculated using the paired t test. D) Changes in the average expression of the immune exhaustion marker *PD-L1* between tumor ROIs from pre-and post-trastuzumab samples in patients classified into the EMT and non-EMT groups. P values were calculated using the paired t test. E) Heatmap of the log2 fold changes in the average expression of inhibitory chemokine genes between tumor ROIs from post-and pre-trastuzumab samples. Rows represent individual chemokine genes, and columns represent patients. Colors indicate the log2 fold change values between post-and pre-trastuzumab samples. Checkpoints that exhibited significant changes in tumors undergoing EMT post-trastuzumab were highlighted. P values were calculated using the paired t test. F) Changes in the average expression of the chemokine gene *CCL2* between tumor ROIs from pre-and post-trastuzumab samples in patients classified into the EMT and non-EMT groups. P values were calculated using the paired t test.

Notably, pathway analysis revealed upregulation of CCR and immune checkpoint gene signatures in the EMT-positive tumors (**Supplementary Figure 4B**). Changes in EMT scores significantly correlated with increased expression of inhibitory immune checkpoints, chemokines, TILs, and CAF-S1 signatures post-trastuzumab (**Figure 4B**). We examined patient-wise changes in inhibitory immune checkpoint expression. EMT-positive tumors exhibited significantly increased expression of 10 checkpoints (P<0.05, paired t-test) and decreased expression of 1 checkpoint post-trastuzumab (**Figure 4C**). In contrast, non-EMT GCs showed no significant differences in any checkpoint gene expression post-treatment. Notably, the immune exhaustion markers *PD-1/PD-L1* and *TIGIT* showed significantly elevated expression in tumors from the EMT group but not the non-EMT group (**Figure 4D**, **Supplementary Figure 4C**). These immune-suppressive changes were further supported by elevated expression of 9 inhibitory chemokines, with 5 showing significant changes (P<0.05, paired t-test) (**Figure 4E**). For instance, *CCL2* and *CCL5*, two chemokines previously reported to be associated with trastuzumab resistance^43,44^, exhibited significantly elevated expression in tumors from the EMT group but not the non-EMT group (**Figure 4F**, **Supplementary Figure 4D**). These findings suggest that tumors in GCs exhibiting EMT post-trastuzumab therapy may represent a distinct molecular subtype with the propensity to upregulate immune checkpoint pathways after molecular therapy.

To independently validate our findings *in vivo*, we utilized patient-derived xenografts (PDXs). Specifically, we established a PDX model (GTR0108) harboring *ERBB2* gene amplification, with HER2 positivity confirmed by IHC/FISH analysis^45^. The HER2+ PDX was treated with trastuzumab until acquired resistance emerged. Real-time PCR analysis revealed that the resistant GTR0108 PDX exhibited elevated *TEAD2* and *ZEB2* gene expression compared to controls, suggesting that EMT pathway activation likely drives resistance in this model (**Supplementary Figure 4E**). Notably, no differences in *PD-L1* expression were observed in the resistant GTR0108 PDX compared to controls, potentially due to the absence of an intact immune system in the PDX model, consistent with previous findings^46^.

### Post-Trastuzumab Tumors not Undergoing EMT may Develop Resistance via the ERAD Pathway

We next investigated the remaining two-thirds of GCs not undergoing EMT but nevertheless exhibited trastuzumab acquired resistance (non-EMT). Differential analysis comparing tumor ROIs before and after treatment in non-EMT patients identified 48 genes with significantly higher post-trastuzumab expression (**Figure 5A**). Pathway analysis revealed a significant increase in the expression of endoplasmic-reticulum-associated protein degradation (ERAD) genes, mTOR signaling, protein secretion pathways, and M2-like macrophage signatures. Conversely, genes associated with IFNα, IFNγ, CCR and immunity-high signatures showed significant expression reduction (**Figure 5B**). ERAD, critical for misfolded protein elimination^47^ and HER2+ breast cancer (BC) cell survival^48^, was the top enriched pathway. ERAD pathway alterations significantly correlated with changes in gene expression related to ER stress, unfolded protein response, *ERBB2* signaling, and PI3K/AKT/mTOR pathways post-trastuzumab treatment (**Figure 5C**). At the level of individual genes, we observed a positive correlation between the expression levels of *RPS13*, *RPL13A*, *GOLM1*, *GRN*, *CTSA*, and ERAD scores in tumor ROIs (**Supplementary Figure 5A**).

**Figure 5.**
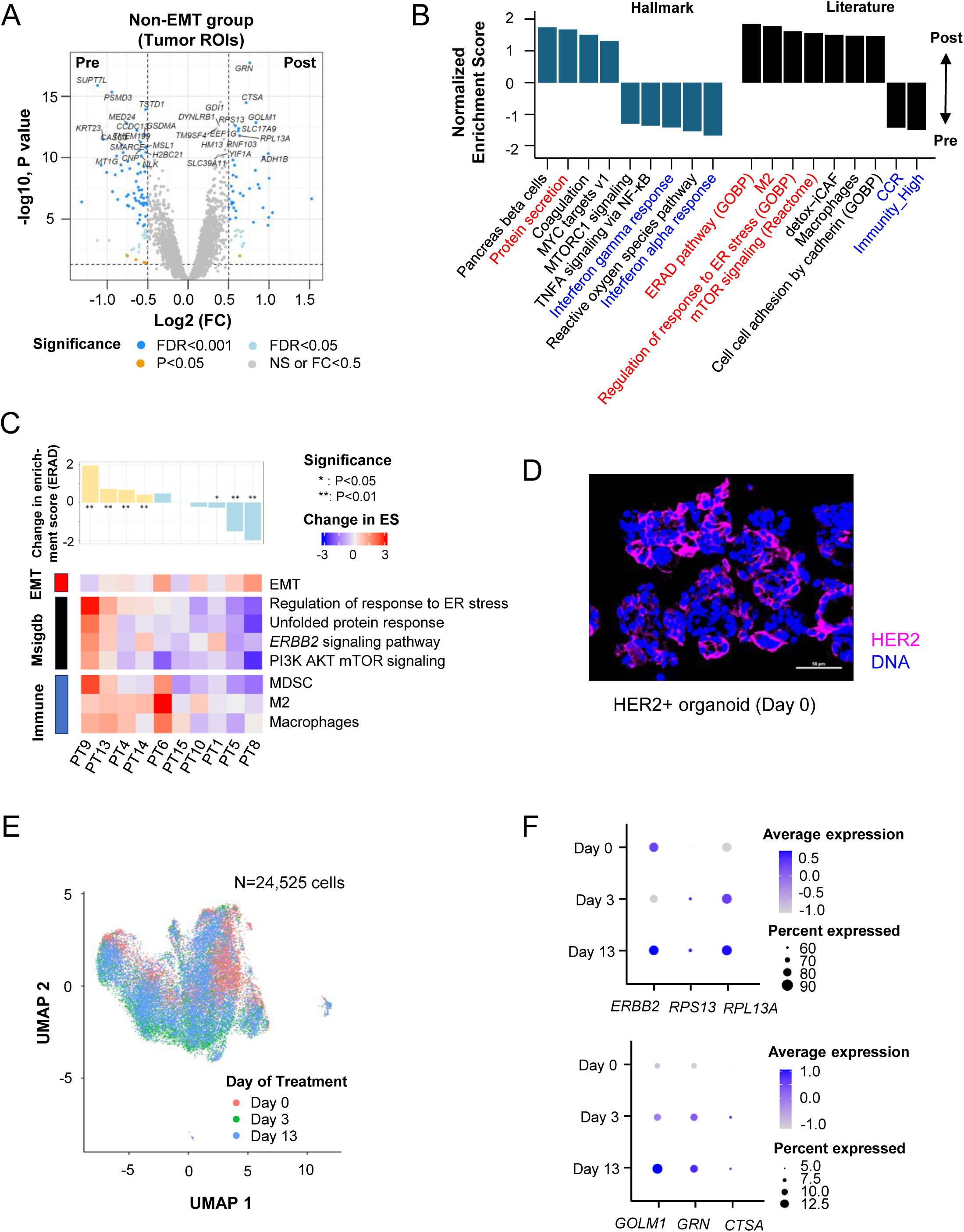
Post-trastuzumab tumors not undergoing EMT have increased expression of ERAD-associated genes. A) Volcano plot comparing gene expression between tumor ROIs from pre-and post-trastuzumab samples of the “non-EMT” patients. The axes represent Log2 FC and significance levels. B) Summary of enrichment in hallmark gene sets and literature gene signatures using differential analysis of expression data between tumor ROIs from pre-and post-trastuzumab samples of the “non-EMT” patients. All gene sets with P<0.05 are shown. C) Change in the average enrichment scores of key differentially regulated gene sets in tumor ROIs from ‘‘non-EMT’’ and ‘‘EMT’’ patient samples (pearson correlation P<0.05). The data are ordered according to the change in enrichment scores of the ERAD pathway. D) Immunofluorescence (IF) staining of the HER2+ organoid on Day 0. E) UMAP visualization of snRNA-seq profiling of HER2+ organoid cells collected on Day 0, Day 3 and Day 13. Cells are colored by the day of treatment. F) Dot plots depicting the expression levels of *ERBB2* and key ERAD-associated genes in the HER2+ organoid at various time points.

To orthogonally corroborate our findings *in vitro*, we employed patient-derived organoids (PDOs). Specifically, we established a PDO with high *ERRB2* expression levels and confirmed HER2 protein-level positivity by Immunofluorescence (IF) staining (**Figure 5D**, **Supplementary Figure 5B**). Interestingly, intratumor HER2 heterogeneity was observed within the PDO. The HER2+ PDO was treated with trastuzumab and subjected to snRNA sequencing at three time points: Day 0, Day 3 and Day 13. UMAP projection and Seurat clustering revealed 12 epithelial subclusters, highlighting the presence of intra-organoid heterogeneity (**Supplementary Figure 5C**). Post-trastuzumab treatment, the transcriptomic profiles shifted notably from Day 0 to Day 13 (**Figure 5E**). Notably, *ERBB2* expression initially decreased in the short term and then increased in the long term, suggesting an initial response followed by acquired resistance (**Figure 5F**). Comparing Day 0 and Day 13, we observed significant increases in the expression of *RPS13*, *RPL13A*, *GOLM1*, *GRN*, and *CTSA*, while EMT signatures decreased, indicating that ERAD pathway activation likely drives resistance in this PDO (**Supplementary Figure 5D**).

### Increases in Oxidative Phosphorylation and HLA Loss may Potentially Drive T-DXd Resistance

Finally, of the 15 patients receiving trastuzumab, 6 were subsequently treated with T-DXd as second-line therapy. Post T-DXd, two patients achieved PR, three exhibited SD, and one had unavailable information. Comparing post-trastuzumab (but prior to T-DXd therapy) tumor ROIs, we identified 110 genes with higher expression in PR patients, including *COL6A1* (**Figure 6A**), and 55 genes with higher expression in SD patients, such as *TACSTD2* encoding the cancer-associated glycoprotein Trop2^49,50^.

**Figure 6.**
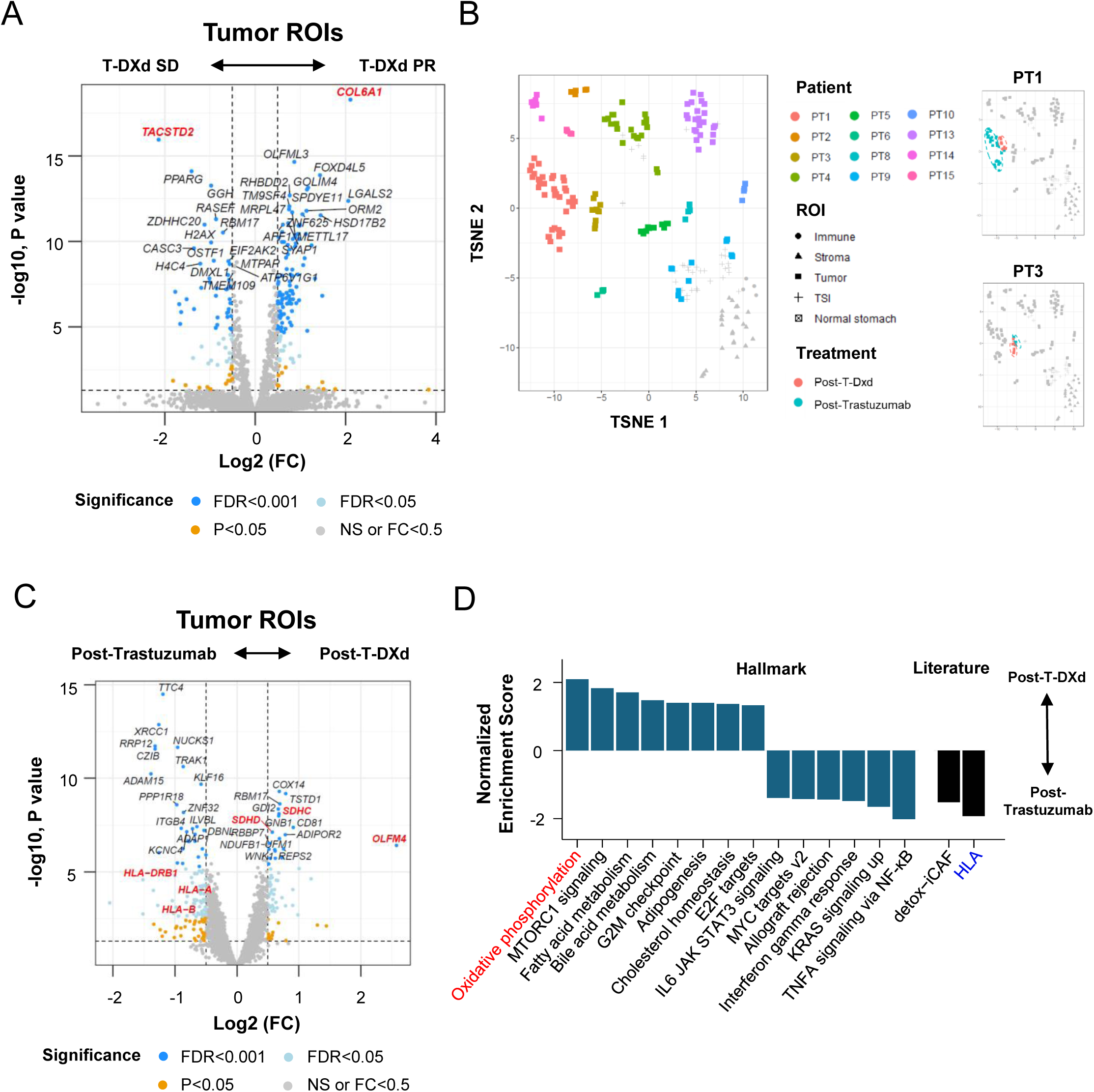
Post-T-DXd tumors show increases in oxidative phosphorylation and HLA loss. A) Volcano plot comparing gene expression between tumor ROIs from the post-trastuzumab samples in patients with SD and PR to T-DXd. The axes represent Log2 FC and significance levels. B) t-SNE plots (left) of ROIs from post-trastuzumab and post-T-DXd samples in HER2+ patients, distinguished by cell type and patient origin. Tumor ROIs are color-coded according to individual patients, whereas non-tumor ROIs are shown in grey. Each point on the t-SNE plot represents a single ROI. t-SNE plots (right) of ROIs from post-trastuzumab and post-T-DXd samples in patients PT1 and PT3, with tumor ROIs color-coded according to the treatment timeline. C) Volcano plot comparing gene expression between tumor ROIs from post-trastuzumab and post-T-DXd samples of the same patient. The axes represent Log2 FC and significance levels. D) Summary of enrichment in hallmark gene sets and literature gene signatures using differential analysis of expression data between tumor ROIs from post-trastuzumab and post-T-DXd samples of the same patient. All gene sets with P<0.05 are shown.

To capture transcriptomic changes profiles following T-DXd acquired resistance, we compared paired post-trastuzumab and post-T-DXd samples from 3 patients. All paired ROIs displayed similar transcriptomic profiles following T-DXd treatment (**Figure 6B**). Differential expression analysis identified 58 genes with higher expression in the tumor ROIs of post-T-DXd samples (**Figure 6C**), including the stem cell maker gene *OLFM4*. Pathway analysis revealed a significant upregulation of oxidative phosphorylation genes, alongside a significant reduction in the expression of genes related to HLA gene signatures upon T-DXd treatment such as *HLA−A* and *HLA-B* (**Figure 6D**). HLA loss has been linked to resistance to immunotherapy and targeted therapy in various cancers^51–53^. These findings suggest HLA loss as a potential mechanism of T-DXd resistance and highlight the role of the immune system in T-DXd’s action.

## Discussion

Our spatial transcriptomics analysis revealed that *ERBB2*-high tumors are characterized by low *PD-L1* expression and lymphocyte infiltration, indicating an ’immune-cold’ TME. However, *PD-L1* expression was higher in stromal regions associated with *ERBB2*-high tumors. This suggests that the conflicting reports on the immune contexture of HER2+ GC may stem from intra-tumor heterogeneity of PD-L1 expression, both within different tumor regions and/or between tumor and non-tumor microregions. Intra-tumoral HER2 heterogeneity, which is prevalent in GC patients and linked to prognosis and trastuzumab response^6–8,54^, was confirmed by our spatial analysis, showing high heterogeneity within HER2+ GCs and consistently low TIL levels in *ERBB2*-high regions. Additionally, our analysis revealed elevated expression of *CDK12* and *ERBB3* in *ERBB2*-high tumors. Upregulation of CDK12 and HER3 is associated with tumorigenesis and trastuzumab resistance^55,56^, and targeting these molecules can reverse trastuzumab resistance in HER2+ cancer *in vivo*^56–58^. Thus, combining therapies targeting HER3 and/or CDK12 with trastuzumab could be a promising strategy for trastuzumab-resistant GC.

EMT contributes to trastuzumab resistance in HER2+ BC and GC^18,19,59,60^. Zhou *et al.* found that the trastuzumab-resistant GC cell line NCI-N87/TR had elevated levels of EMT TFs, including *ZEB1* and *ZEB2*^19^. Our study observed EMT development in about one-third of GCs with acquired trastuzumab resistance, with upregulated immune checkpoints such as *PD-1/PD-L1* post-treatment. Multiple studies have shown a strong positive correlation between EMT status and immune checkpoint expression, especially *PD-L1* expression, in various cancers^61–63^. The induction of PD-L1 after trastuzumab therapy suggests that chemotherapy and targeted therapy may modify the TME, offering potential synergy in combining HER2 and PD-L1 blockade, even in initially immune-cold tumors. It also highlights the potential of combined HER2 and EMT blockade.

In our study, trastuzumab resistance was also associated with the upregulation of *CLDN18* alongside increased TIL and immunity-high signatures. The human *CLDN18* gene produces the CLDN18.2 protein isoform, and CLDN18.2 has been identified as a therapeutic target^41,64,65^. The efficacy of anti-CLDN18 antibody drugs in patients with GC has been demonstrated^41,66,67^. Notably, recent findings have demonstrated that *CLDN18* expression enhances T lymphocyte infiltration and antitumor immunity in pancreatic cancer^68^. Thus, similar to targeting both PD-1/PD-L1 and HER2, a dual targeting approach of HER2 and CLDN18.2 may offer a promising strategy to overcome trastuzumab resistance.

Our study also highlights the ERAD pathway’s role in trastuzumab resistance in HER2+ GC. Increased expression of ERAD pathway genes has been observed in HER2+ BC^48^. Inhibition of ERAD induces irrecoverable ER stress and selectively eradicates HER2+ BC cells. In our study, non-EMT tumors exhibited upregulation of ERAD, mTOR signaling, and protein secretion pathways. The upregulation of mTOR signaling and protein secretion pathways may indicate an accumulation of proteotoxic burden within tumor cells, which is alleviated by increased ERAD activity. Our findings propose that targeting ERAD pathways alongside immunotherapy could be a promising strategy for trastuzumab-resistant GC.

Finally, T-DXd therapy has significantly improved response and overall survival in HER2+ GC and BC^11,69^, but resistance remains common. Reductions in HER2 expression and loss-of-function mutations in *SLX4* have been identified as candidate resistance mechanisms to T-DXd in BC^70^. However, mechanisms of T-DXd resistance in GC remain largely unexplored. Interestingly, we observed transcriptional loss of HLA and upregulation of oxidative phosphorylation following T-DXd treatment. T-DXd incorporates an IgG1 backbone, enabling antibody-dependent cellular cytotoxicity (ADCC) *in vivo*, thereby retaining the immune-attracting properties of the naked monoclonal antibody and functioning as a form of immunotherapy^71^. Disruption of HLA Class I antigen processing mediates escape from immune checkpoint inhibitors (ICIs) in multiple cancers^51,52^. Inhibiting oxidative phosphorylation has reversed resistance to HER2-targeted therapies in HER2+ BC and may be a potential strategy for T-DXd-resistant GC^72^. A potential limitation of our study is the small number of T-DXd-treated patients included for transcriptome profiling.

In summary, our spatial profiling of HER2+ GC identified *ERBB2*-high tumor regions exhibiting immune-cold phenotypes both within and across patients. Additionally, we delineated two distinct mechanisms of acquired resistance to trastuzumab in HER2+ GC, associated with the onset of EMT or ERAD pathway induction. Our study also revealed potential therapeutic targets, such as *PD-L1* and *CLDN18*, for treating trastuzumab-resistant patients. Furthermore, we identified tumor HLA loss as a potential mechanism of T-DXd resistance. Overall, these findings can guide more rational therapeutic strategies to prevent, overcome, and reverse acquired resistance to anti-HER2 treatment in patients with HER2+ GC.

## Data Availability

All data produced in the present work are contained in the manuscript.

## Acknowledgements

The authors would like to thank Monia Niero from Department of Pathology, Azienda ULSS 2 Marca Trevigiana, Treviso, Italy for the technical support.

