## Supplementary Figures for "Spatial Profiling of Patient-Matched HER2 Positive Gastric Cancer Reveals Resistance Mechanisms to Trastuzumab and Trastuzumab Deruxtecan"

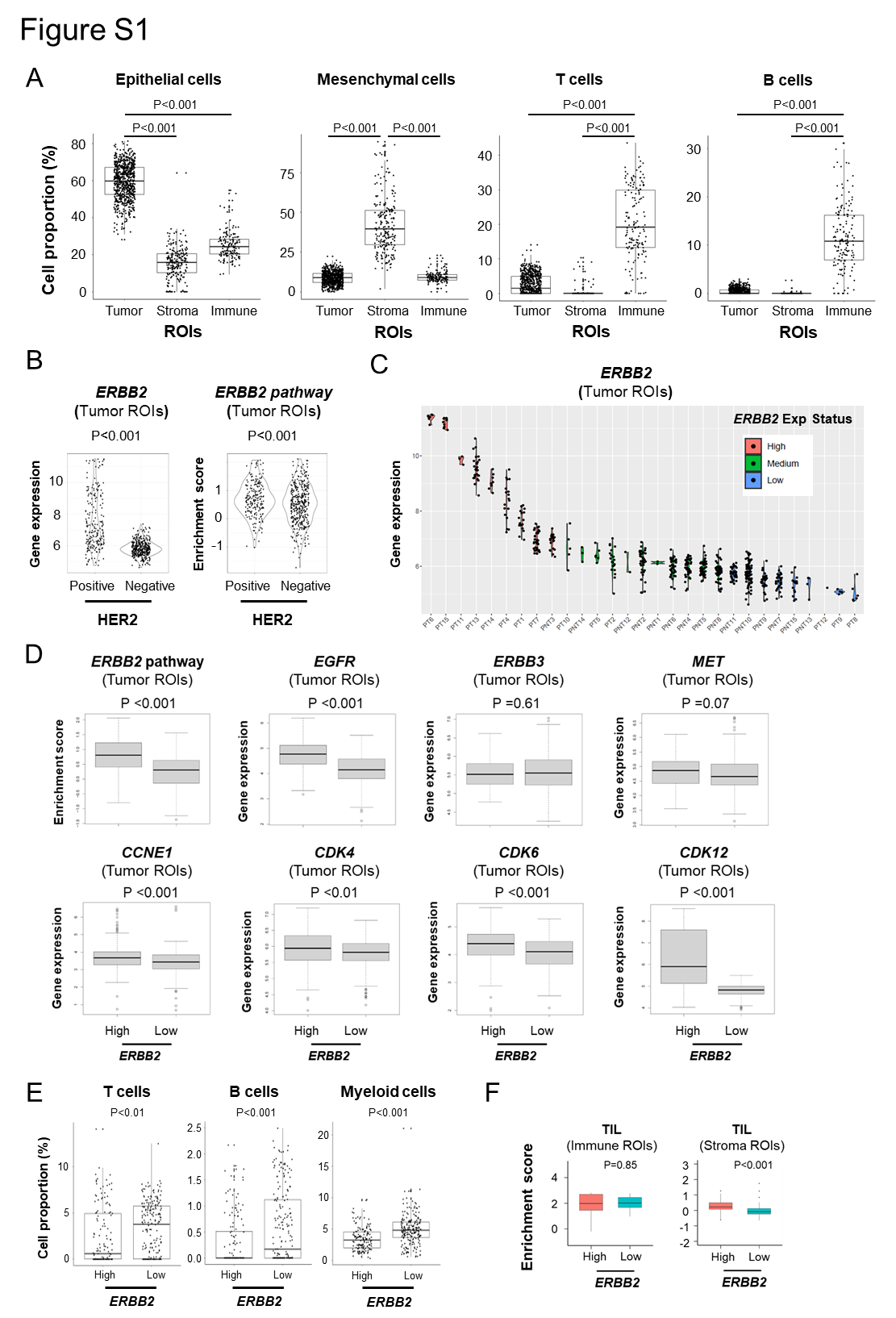

**Supplementary Figure 1**

A) Comparison of the proportions of deconvoluted cell types among tumor, stroma, and immune regions of interest (ROIs) from samples across all three sub-cohorts. P values were calculated using the Wilcoxon test.
B) Difference in *ERBB2* expression levels and *ERBB2* pathway enrichment scores in tumor ROIs from pre-trastuzumab samples between HER2+ and HER2-negtaive GCs. P values were calculated using the Wilcoxon test.
C) Violin plot of *ERBB2* expression levels in tumor ROIs from pre-trastuzumab samples. Patients are ordered according to the average *ERBB2* expression levels of tumor ROIs. Tumors are categorized into *ERBB2*-high, *ERBB2*-medium, and *ERBB2*-low groups in equal proportions.
D) Difference in *ERBB2* pathway enrichment scores and expression levels of RTKs and cell cycle genes in tumor ROIs from pre-trastuzumab samples between *ERBB2*-high and *ERBB2*-low GCs. P values were calculated using the Wilcoxon test.
E) Comparison of the proportions of deconvoluted immune cell types in tumor ROIs from pre-trastuzumab samples between *ERBB2*-high and *ERBB2*-low GCs. P values were calculated using the Wilcoxon test.
F) Comparison of TIL enrichment scores of immune and stroma ROIs between *ERBB2*-high and *ERBB2*-low GCs. P values were calculated using the Wilcoxon test.

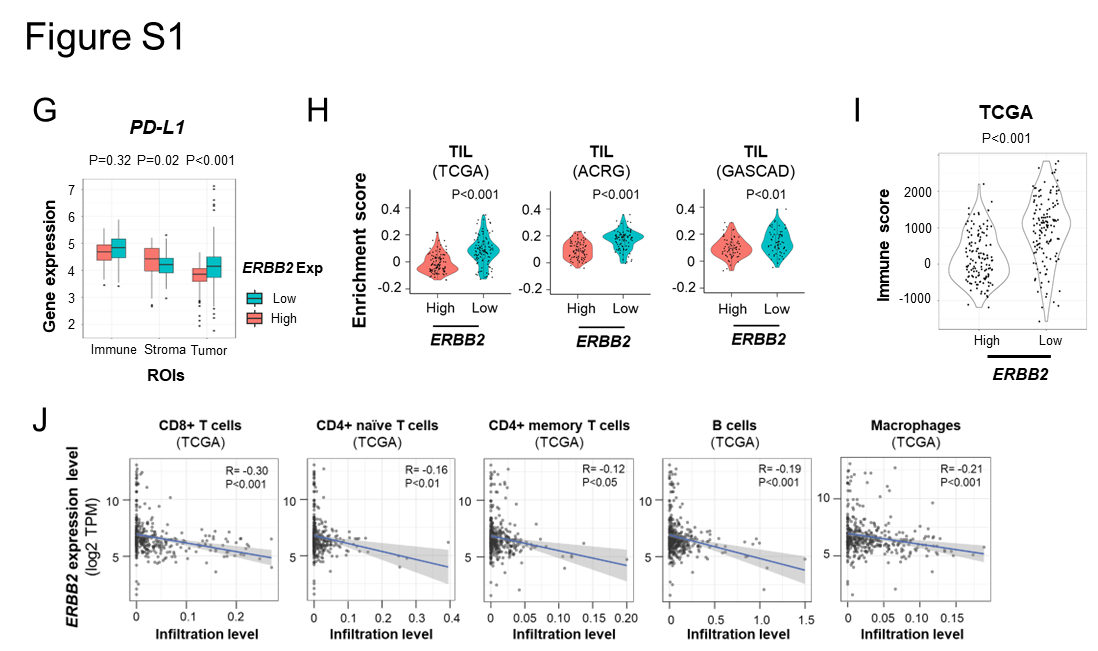

**Supplementary Figure 1**

G) Comparison of the *PD-L1* expression levels among tumor, stroma, and immune ROIs from pre-trastuzumab samples between *ERBB2*-high and *ERBB2*-low GCs. P values were calculated using the Wilcoxon test.
H) Comparison of TIL enrichment scores between *ERBB2*-high and *ERBB2*-low tumors from the TCGA, ACRG and GASCAD GC cohorts. P values were calculated using the Wilcoxon test.
I) Difference in estimated immune scores between *ERBB2*-high and *ERBB2*-low tumors from the TCGA GC cohort. P values were calculated using the Wilcoxon test.
J) Association between *ERBB2* expression levels and immune cell infiltration in tumors from the TCGA GC cohort.

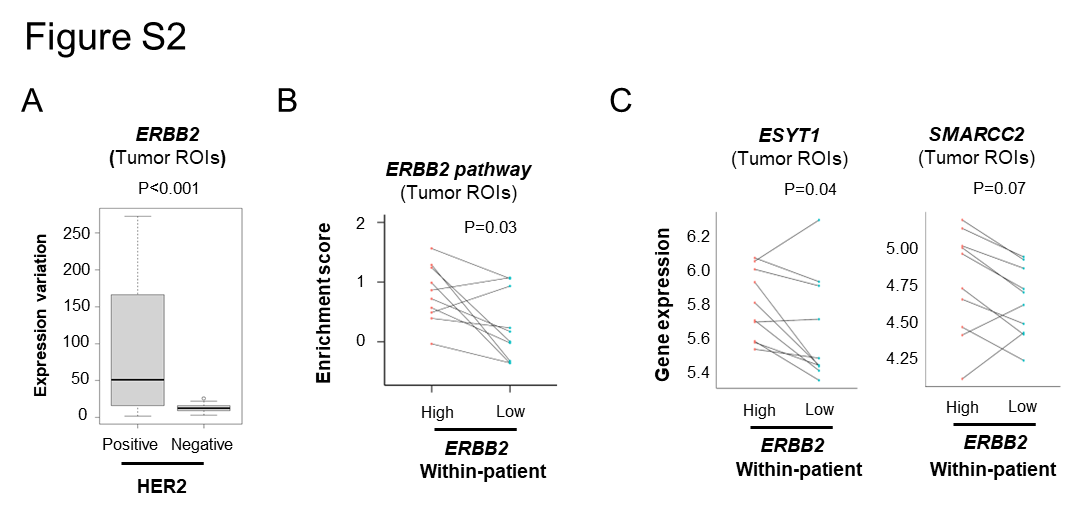

**Supplementary Figure 2**

A) Difference in the variation of *ERBB2* expression in tumor ROIs between HER2+ and HER2-negative GCs. P values were calculated using the Wilcoxon test.
B) Difference in *ERBB2* pathway enrichment scores between *ERBB2*-high and *ERBB2*-low tumor ROIs from the same patient. P values were calculated using the paired t test.
C) Difference in expression levels of *ESYT1* and *SMARCC2* between *ERBB2*-high and *ERBB2*-low tumor ROIs from the same patient. P values were calculated using the paired t test.

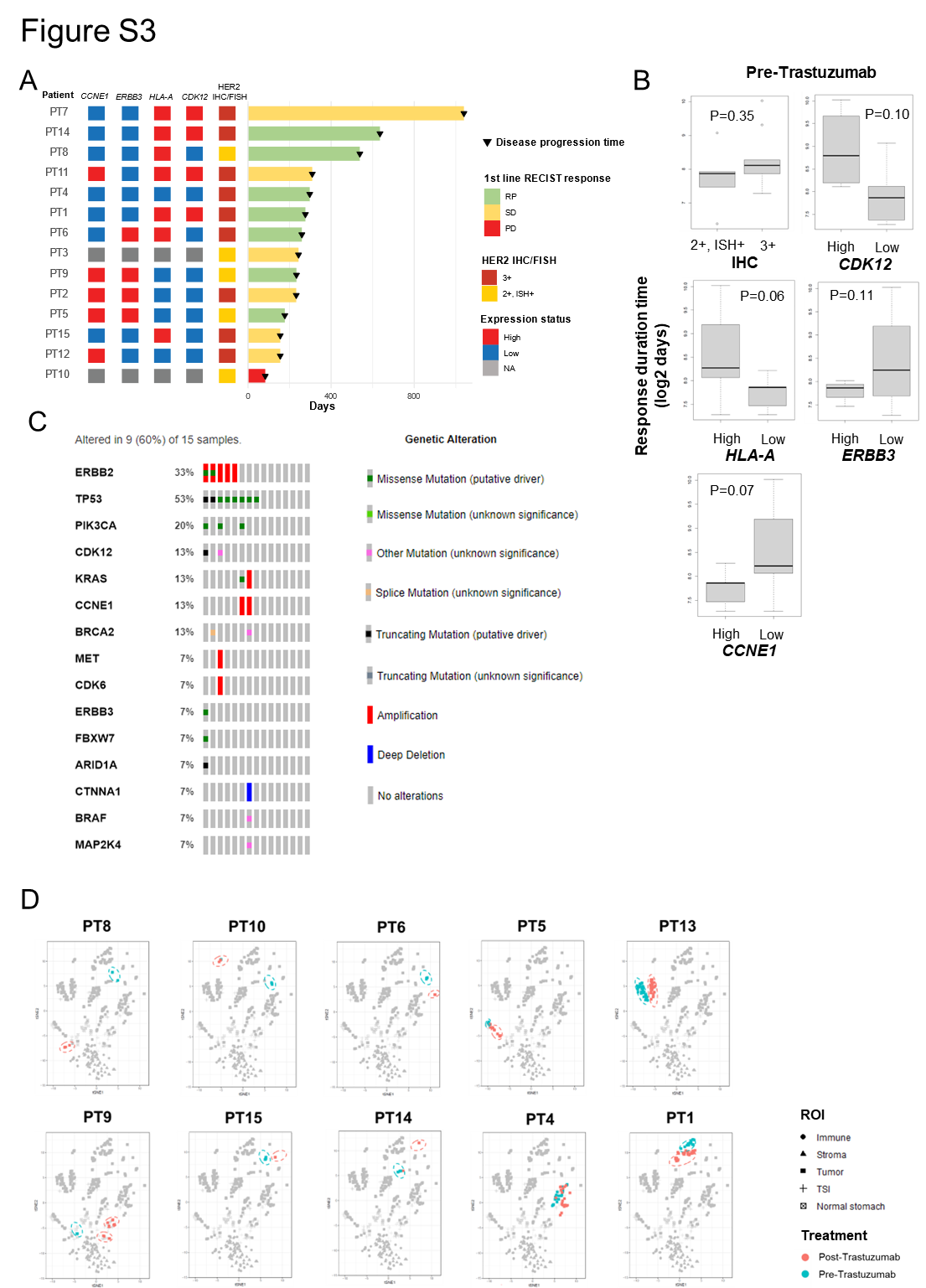

**Supplementary Figure 3**

A) Swimmer's plot of when each patient showed disease progression. The plot includes first-line RECIST response, HER2 IHC/FISH status, and gene expression status. Gene expression status (high or low) was determined by the average expression levels of the corresponding genes in tumor ROIs from the patient's pre-trastuzumab samples.
B) Comparison of response duration time to trastuzumab between HER2+ GCs with an IHC score of 2/ISH+ and those with an IHC score of 3. Additionally, comparison of response duration time to trastuzumab between HER2+ GCs with high and low expression levels of *CDK12*, *HLA-A*, *ERBB3*, and *CCNE1*. P values were calculated using the Wilcoxon test.
C) Genetic alterations in post-trastuzumab samples from 9 HER2+ patients. Top genetic alterations are shown.
D) t-SNE plots of ROIs from pre- and post-trastuzumab samples in HER2+ GCs, with tumor ROIs color-coded according to the treatment timeline.

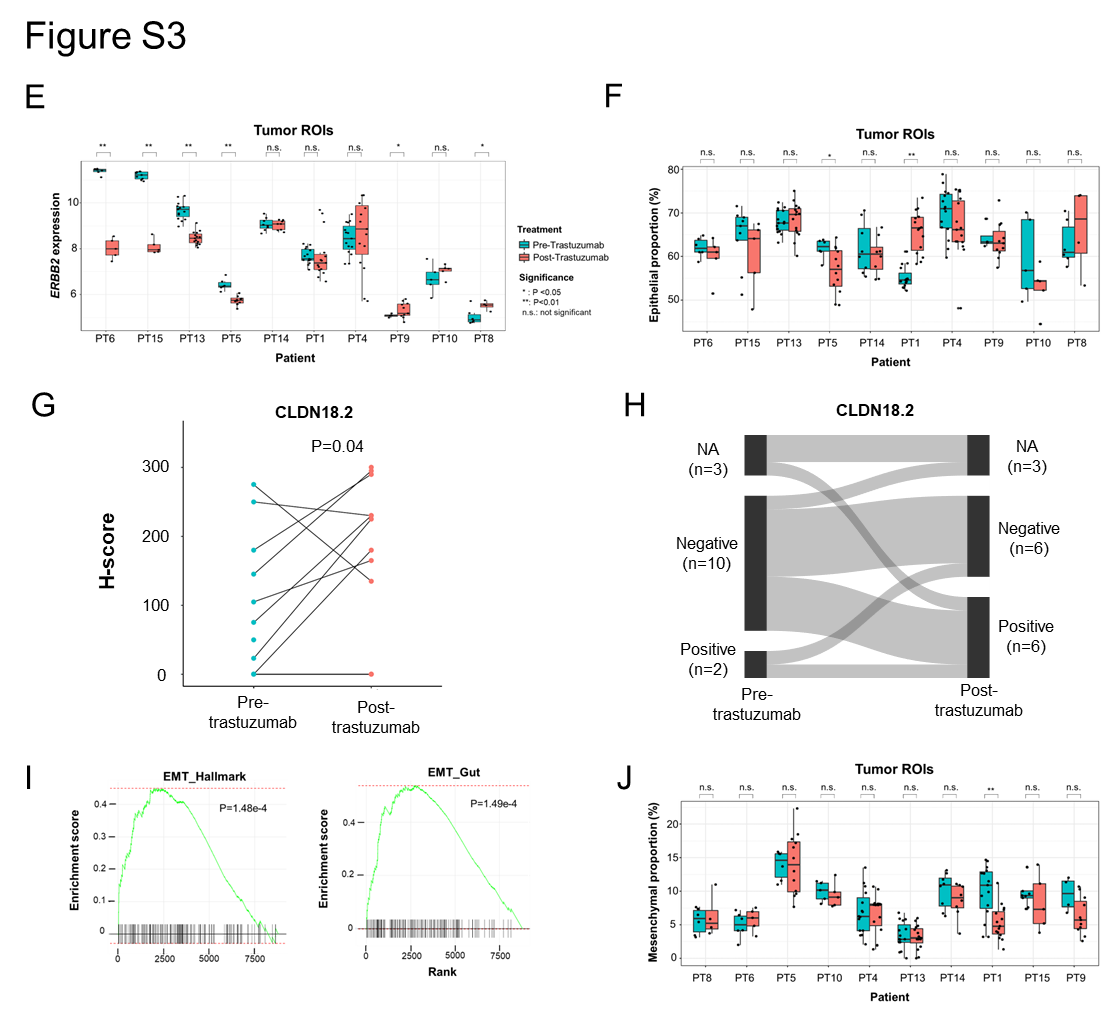

**Supplementary Figure 3**

E) Comparison of *ERBB2* expression levels between tumor ROIs from pre- and post-trastuzumab samples in each patient. Patients are ordered according to the changes in average *ERBB2* expression levels of tumor ROIs. P values were calculated using the Wilcoxon test.
F) Comparison of epithelial proportions between tumor ROIs from pre- and post-trastuzumab samples in each patient. Patients are ordered according to the changes in average *ERBB2* expression levels of tumor ROIs. P values were calculated using the Wilcoxon test.
G) Difference in CLDN18.2 H-scores between pre- and post- trastuzumab samples from the same patient. P values were derived from the linear mixed model.
H) Sankey plot illustrating the changes in CLDN18.2 positivity between pre- and post-trastuzumab samples in HER2+ patients.
I) GSEA analysis of the Hallmark EMT pathway and a published GC EMT gene set using differential analysis of expression data between tumor ROIs from pre- and post-trastuzumab samples of the same patient. P values were generated using GSEA.
J) Comparison of mesenchymal proportions between tumor ROIs from pre- and post-trastuzumab samples in each patient. Patients are ordered according to the changes in average EMT scores of tumor ROIs. P values were calculated using the Wilcoxon test.

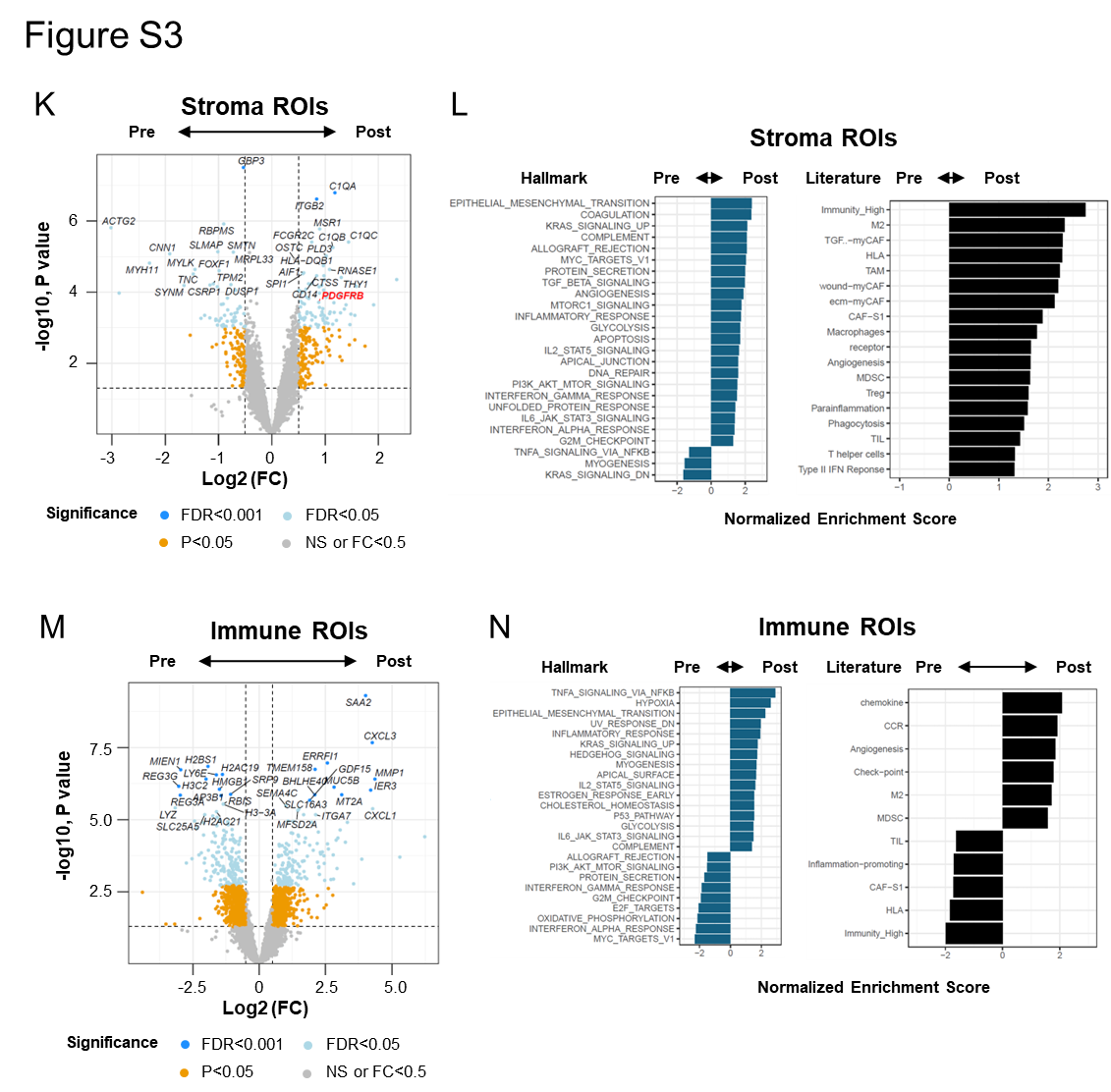

**Supplementary Figure 3**

K) Volcano plot comparing gene expression between stroma ROIs from pre- and post-trastuzumab samples of the same patient. The axes represent Log2 FC and significance levels.
L) Summary of enrichment in hallmark gene sets and literature gene signatures using differential analysis of expression data between stroma ROIs from pre- and post-trastuzumab samples of the same patient. All gene sets with P<0.05 are shown.
M) Volcano plot comparing gene expression between immune ROIs from pre- and post-trastuzumab samples of the same patient. The axes represent Log2 FC and significance levels.
N) Summary of enrichment in hallmark gene sets and literature gene signatures using differential analysis of expression data between immune ROIs from pre- and post-trastuzumab samples of the same patient. All gene sets with P<0.05 are shown.

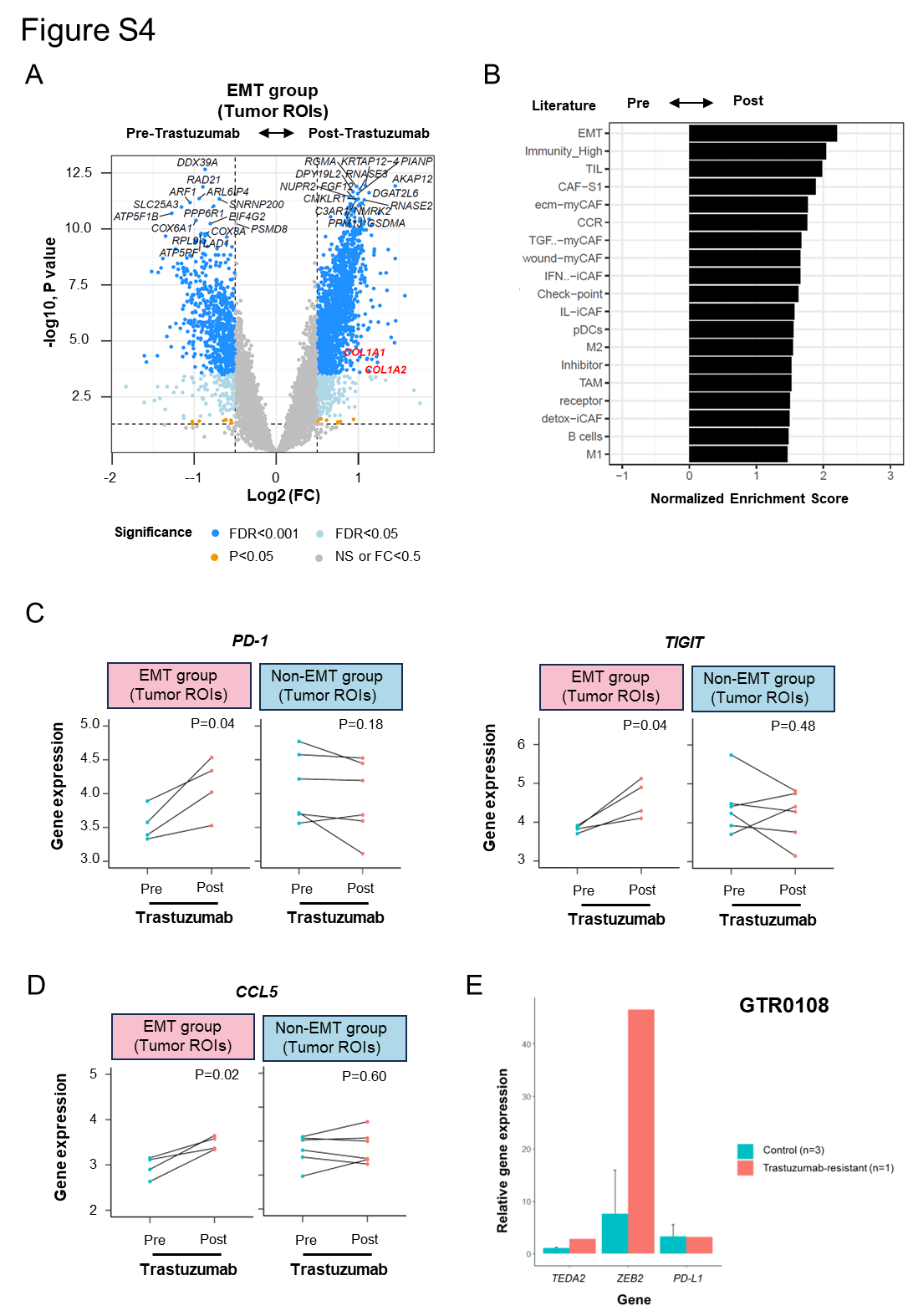

**Supplementary Figure 4**

A) Volcano plot comparing gene expression between tumor ROIs from pre- and post-trastuzumab samples of the “EMT” patients. The axes represent Log2 FC and significance levels.
B) Summary of enrichment in literature gene signatures using differential analysis of expression data between tumor ROIs from pre- and post-trastuzumab samples of the “EMT” patients. All gene sets with P<0.05 are shown.
C) Changes in the average expression of the immune exhaustion markers *PD-1*/*TIGIT* between tumor ROIs from pre- and post-trastuzumab samples in patients classified into the EMT and non-EMT groups. P values were calculated using the paired t test.
D) Changes in the average expression of the chemokine gene *CCL5* between tumor ROIs from pre- and post-trastuzumab samples in patients classified into the EMT and non-EMT groups. P values were calculated using the paired t test.
E) Differences in the gene expression of *TEAD2*, *ZEB2* and *PD-L1* between the resistant GTR0108 PDX and controls.

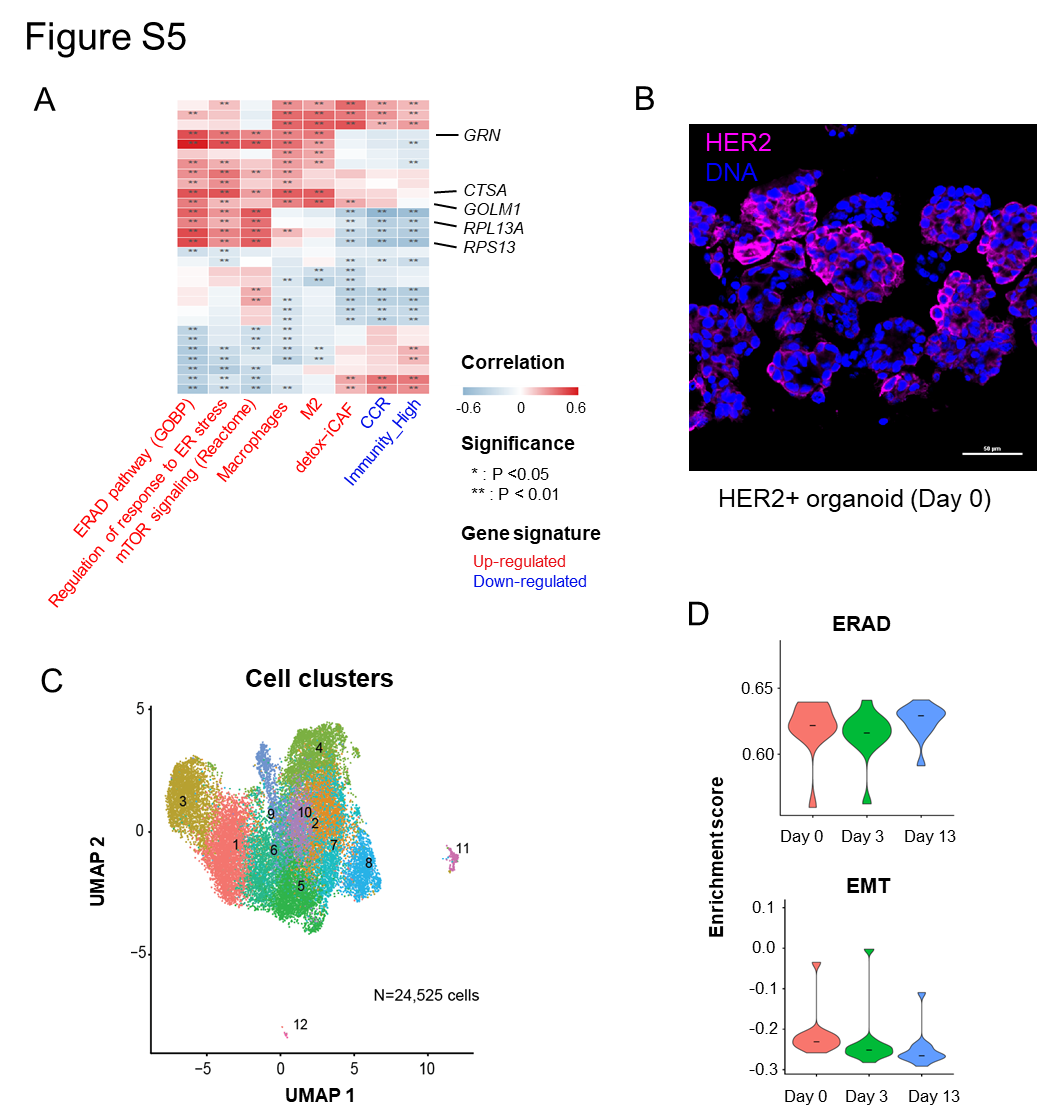

**Supplementary Figure 5**

A) Heatmap showing the correlation between the expression of differential genes and the enrichment scores of differential gene sets in tumor ROIs. Rows represent individual differential genes, and columns represent differential gene sets. Colors indicate the Pearson’s correlation coefficients. P value was calculated by Pearson’s correlation test.
B) Immunofluorescence (IF) staining of the HER2+ organoid on Day 0.
C) UMAP visualization of snRNA-seq profiling of HER2+ organoid cells collected on Day 0, Day 3 and Day 13. Cells are colored by the Cell cluster.
D) Violin plot showing enrichment scores for ERAD and EMT pathways in the HER2+ organoid at different time points.
