## Supplementary Methods for "Spatial Profiling of Patient-Matched HER2 Positive Gastric Cancer Reveals Resistance Mechanisms to Trastuzumab and Trastuzumab Deruxtecan"

**Patient sample collection**

Formalin-fixed, paraffin-embedded (FFPE) tissue samples were obtained from patients (PT1-15) diagnosed with HER2-positive metastatic gastric cancer (GC) who received anti-HER2 standard therapies at the Fondazione IRCCS Istituto Nazionale dei Tumori, Milan, Italy. Archival tumor tissue from surgery or diagnostic biopsies obtained before trastuzumab-based therapy, coupled with a post-progression tumor sample obtained after progression on trastuzumab, was required. An additional tumor re-biopsy obtained after progression on trastuzumab deruxtecan was optional. All experimental protocols adhered to the pertinent legal and institutional regulations and received approval from the respective local ethics committees. Informed written consent was obtained from all participants prior to their inclusion in the study INT 117/15. FFPE tissue samples were also obtained from patients (PNT1-15) diagnosed with gastric adenocarcinoma undergoing surgical resection or endoscopy at the National University Hospital, Singapore. On-table endoscopic biopsies or surgical resection samples were collected along with matched normal gastric tissues from areas several centimeters away from the tumor site. This study was approved by the local ethics board (National Healthcare Group Domain Specific Review Board Ref Nos: 2005/00440 and 2016/00059).

**HER2 immunohistochemistry/in situ hybridization**

HER2 positivity was defined as either a score 3+ at immunohistochemistry (IHC) or a IHC 2+ score and positive in situ hybridization (FISH), by using PATHWAY anti-HER2/neu (4B5) IHC and the INFORM HER2 Dual ISH assays (Ventana Medical Systems), according to standard guidelines^1^.

**CLDN18 immunohistochemistry**

IHC stains were centrally performed on four-micrometer thick FFPE sections using BenchMark ULTRA IHC/ISH System (Ventana) for the staining of CLDN18 (clone 43-14A; Roche Ventana), according to the laboratory’s routine practice and the manufacturer’s instructions. Both the intensity of tumor cell membrane staining and the percentage of cancer cells with complete, basolateral or lateral membrane staining were assessed. CLDN18 positivity was defined as tumor cells showing moderate-to-strong (2+/3+ score) membranous reactivity in ≥75 % of tumor cells^2^. Cytoplasmic or granular staining patterns were disregarded. We also graded both intensity (0, 1+, 2+, 3+) and percentage (0–100 %) of CLDN18 staining and calculated the histoscore (H-score) by multiplying the intensity score with the percentage^3^.

**Foundation One panel**

Comprehensive genomic profiling (CGP) was performed using formalin-fixed paraffin-embedded tumor tissue by Foundation Medicine, Inc, Cambridge, MA, through FoundationOne CDx^4^.

**Digital Spatial Profiling**

FFPE tissues were mounted on Leica Bond slides for the NanoString GeoMx DSP platform. For H&E staining, FFPE slides were deparaffinized with histoclear, rehydrated and stained with Hematoxylin Solution. Slides were counterstained with eosin and mounted with mounting media. FFPE slides were subjected to conventional tissue preprocessing (deparaffinization and rehydration). Standard fluorescence-labeled morphology marker panel consisting of Pan-CK for epithelial regions, CD45 for immune cells, α-smooth muscle actin for fibroblast and nuclear stain were used as Regions of interest (ROIs) selection references. ROIs for each slide were drawn and selected.

**DSP Data Processing and Analysis**

FASTQ files from Digital Spatial Profiling (DSP) were processed into count matrices following established protocols^5^. Sequencing counts were deduplicated based on unique molecular identifiers (UMIs) and molecular target tag sequences, with single-probe genes reported as deduplicated count values. Data processing and normalization were conducted using the GeoMxTools R package v2.0. ROIs that failed to meet quality criteria—defined as having fewer than 1,000 raw reads, less than 75% read alignment, or sequencing saturation below 50%—were excluded from further analysis. The quantitation limit was set at two geometric standard deviations above the geometric mean of the negative control probes. ROIs with fewer than 5% of panel genes exceeding this quantitation limit, as well as genes detected in less than 10% of the remaining ROIs, were removed. Data normalization was achieved through upper quartile (Q3) normalization. Batch correction was undertaken with RUV4^6^. Following filtering, each ROI exhibited a median of more than 3,600 detected genes, with the interquartile range (IQR) detailed subsequently. Overall, the ROIs accounted for a comprehensive profile of detected genes. Dimension reduction of DSP data was conducted with *t*-distributed Stochastic Neighbor Embedding (TSNE) using the R package Rtsne (v.0.15). Cell abundances within each ROI were estimated using the SpatialDecon algorithm (v.1.4.3), leveraging a human cell-profile reference matrix available at Nanostring Biostats GitHub <https://github.com/Nanostring-Biostats/CellProfileLibrary/tree/master/Human>^7^.

Differential expression analysis was performed using a linear mixed-effect model (LMM). When comparing paired samples from different patients (eg pre-trastuzumab vs post-trastuzumab), a random Intercept on patient is included in the LMM model. Gene Set Enrichment Analysis (GSEA) of the differentially expressed genes was conducted using a comprehensive, non-redundant resource of gene sets, including Hallmark gene sets (<https://www.gsea-msigdb.org/gsea/msigdb/>) and literature gene sets from bulk or single-cell RNA-seq through the R fgsea package (v.1.20.0). Enrichment scores for gene signatures and pathways in the DSP data were calculated via single-sample gene set enrichment analysis (ssGSEA) using the GSVA package (v.1.42.0) in R.

**Transcriptomic data analysis of public GC cohorts**

Level 3 TCGA RNA-seq normalized matrix for 415 GC gastric samples was downloaded from the Broad Institute TCGA Genome Data Analysis Center (GDAC) Firehose (<https://gdac.broadinstitute.org/>). Gene expression of 300 GC patients in the ACRG cohort was profiled using Affymetrix Human Genome U133plus 2.0 Array (GSE62254: <https://www.ncbi.nlm.nih.gov/geo/query/acc.cgi?acc=GSE62254>)

and processed as described previously^8^. Gene expression of primary GC tumors in the GASCAD cohort was profiled using Affymetrix GeneChip Human Genome U133 Plus 2.0 Array (GSE15459: <https://www.ncbi.nlm.nih.gov/geo/query/acc.cgi?acc=GSE15459>) and processed as described as previously^9^. Enrichment scores for gene signatures and pathways in the transcriptomic data of tumors from the three GC cohorts were determined using ssGSEA with the GSVA package in R. The infiltration level of immune cells in GC tumors from the TCGA cohort based on gene expression data were analyzed by ESTIMATE^10^. The association between infiltration levels of various immune cell types and *ERBB2* expression in GC tumors from the TCGA cohort, as estimated by the XCELL immune deconvolution method, was analyzed using the TIMER2.0 web server^11^.

**Generation of gastric cancer PDXs with acquired resistance to trastuzumab**

Gastric PDX generation was performed as previously described^12^. All animal procedures adhered to the ‘Animal Research: Reporting of In Vivo Experiments’ (ARRIVE) standards and were approved by the Ethical Committee of the Candiolo Cancer Institute, and by the Italian Ministry of Health.

GTR0108 PDXs^13^, bearing *ERBB2* gene amplification, were passaged and expanded for >2 generations until production of a cohort of mice. Established and randomized tumors (average volume 250 mm^3^) were treated with trastuzumab 30 mg/Kg, weekly i.p., until the onset of secondary resistance (i.e. exponential tumor growth in the presence of the treatment). Vehicle (saline) treated mice were used as control. Tumor size was evaluated once-weekly by caliper measurements and approximate volume of the mass was calculated using the formula 4/3π(D/2)(d/2)^2^, where D is the major tumor axis and d is the minor tumor axis.

**Gene expression analysis by real-time PCR**

Total RNA from tumors was isolated with the Maxwell RSC miRNA Tissue Kit (AS1460, Promega), according to the manufacturer’s instructions. The cDNA preparation was done according to standard procedures, using High Capacity cDNA reverse transcription kit (4368814, Applied Biosystem), with RNAase inhibitor (N8080119, Thermo Fisher Scientific). Gene expression was measured by means of the following TaqMan probes (Thermo Fisher Scientific): PD-L1: (CDC274) Hs00204257_m1; ZEB2: Hs00207691_m1; TEAD2: Hs01055894_m1.

**Establishment of Organoids and Identification of HER2+ Organoids**

Patients at The National University Hospital in Singapore were enrolled in this study following approval from the local ethics board (National Healthcare Group, Domain Specific Review Board Ref No: 2005/00440). Specimens were obtained via endoscopic biopsy. Organoids were established using a protocol adapted from Nanki et al.^14^ with some modifications. Briefly, tissues were thoroughly washed with phosphate-buffered saline (PBS) and mechanically minced using a scalpel. Subsequently, enzymatic digestion was performed with Liberase TH (Cat#05401151001, Roche) at 37 °C for 10-30 minutes, following by treatment with TrypLE Express (Cat#12563011, Gibco) at 37 °C for 15 minutes for undigested pellets. Dissociated cells were washed with PBS supplemented with 10% fetal bovine serum (FBS) to inactivate digestive enzymes. The cells were embedded in Matrigel Growth Factor Reduced (Cat#356231, Corning), overlaid with complete organoid medium, and cultured in 5% CO2 and 20% O2. The culture medium was refreshed every 2 to 3 days. The basal medium consisted of Advanced Dulbecco’s Modified Eagle’s Medium/F12(Cat#12634010, Gibco) was supplemented with penicillin/streptomycin, 50% WNT3A conditioned medium, 10% RSPO-1 conditioned medium, 10mM HEPES (Cat#15630080, Gibco), 2mM GlutaMAX (Cat#35050061, Gibco), 1 x B27 (Cat#17504044, Gibco), and 1 mM N-acetylcysteine (Cat#A9165-5G, Sigma-Aldrich). Additional growth factors and small molecules were added to the basal medium to prepare the complete medium, including 50 ng/ml mouse recombinant EGF (Cat#PMG8043,Gibco), 100 ng/ml human recombinant FGF10 (Cat#100-26-250UG, Peprotech), 100 ng/ml mouse recombinant noggin (Cat#250-38-250, Peprotech), 1 nM gastrin I (Cat#G9145-.1MG, Sigma-Aldrich), 2 uM A83-01 (Cat#2939, Tocris), and 10uM Y-27632 (Cat#Y0503-5MG, Sigma-Aldrich). To prevent mycoplasma contamination, 100ug/ml Primocin (Cat#ant-pm-1, InvivoGen) was added during the first 2 weeks. For cancer organoid enrichment, manual selection was performed under microscopy in a subset of cancer organoid lines. Organoids passaged at least 5 times were confirmed to be free of mycoplasma contamination using the MycoAlert PLUS Mycoplasma Detection kit (Cat#LT07-703, Lonza), preserved as frozen stocks with CryoStor cell cryopreservation media, CS10 (Cat#C2874-100ML, Sigma-Aldrich), and their recovery after freeze-thawing process was assessed. Whole exome sequencing (WES) and Whole Transcriptome sequencing (WTS) were performed on fully-recovered organoids post-thawing for organoid profiling. Based on CNV analysis and HER2 expression analysis, organoids were identified as HER2 positive organoids.

**Immunofluorescence staining for HER2 in Organoids**

Immunofluorescence (IF) staining was performed to evaluate the expression of HER2 in HER2-positive organoids using an anti-HER2 antibody (Cat#4290S, Cell Signaling TECHNOLOGY). Briefly, 7,500 organoids were recovered from Matrigel using Cell Recovery Solution (Cat#354253, Corning) and embedded in iPGell (Cat#FNK-PG20-1, GenoStaff) according to the manufacturer's instructions. The organoids were then fixed in 10% neutral buffered formalin, embedded in paraffin, and 5 µm FFPE (formalin-fixed, paraffin-embedded) sections were prepared. Antigen retrieval was performed by incubating the sections in a 97°C water bath for 10 minutes with either pH 6 (citrate) or pH 9 (tris) buffer. The sections were incubated with the primary antibody at 4°C overnight, followed by incubation with the secondary antibody at room temperature for one hour. Finally, the sections were stained with DAPI (Cat#130-111-570, Miltenyi Biotec) for one minute before observation.

**Organoid Sample Preparation and Sequencing**

Organoids were treated with 1 µM Trastuzumab (Cat#A2007, Selleckchem). Organoids were collected at three time points: before treatment (day 0), after short-term treatment (day 3), and after long-term treatment (day 13). Nuclei were isolated from the organoids using Nuclei Extraction Buffer (Cat#130-128-024, Miltenyi Biotec) and the gentleMACS Octo Dissociators (Miltenyi Biotec) according to the MACS recommended protocol. The nuclei were passed through 30 µm MACS SmartStrainers (Cat#130-098-458, Miltenyi Biotec) and counted using a manual hemocytometer, targeting 7,500 nuclei per sample. Library preparation was performed using the 10x Genomics kit following the manufacturer's instructions (CG000338, Rev. A). The GEX libraries were sequenced on a NovaSeq X Plus (PE150).

Cellranger v3.0 ([**https://support.10xgenomics.com/single-cell-gene-expression/software/**](https://support.10xgenomics.com/single-cell-gene-expression/software/)) was used to align FASTQ sequencing reads to the hg38 reference transcriptome, generating single-cell feature counts for each sample. Utilizing Seurat version 4.0^15^, each sample was analyzed for genes/features present in three or more cells, and cells with 500 or more features. The single-cell datasets were subsequently combined. Cells with mitochondrial RNA percentages exceeding 17%, fewer than 500 features, or more than 7,000 features were excluded. SCTransform normalization was applied, with mitochondrial RNA included as a regression variable. The integrated data were scaled and subjected to principal component analysis. Visualization was achieved using Uniform Manifold Approximation and Projection (UMAP). Cell clusters were delineated using a shared nearest-neighbor (SNN) modularity optimization-based clustering algorithm at a resolution of 0.8.

**Statistical Analysis**

Statistical analyses were conducted using the R software (version 4.1.2). Significance was determined with a threshold set at p<0.05, adjusted for multiple testing where applicable. Continuous variables, such as gene expression and cell proportion, were assessed using the Wilcoxon rank-sum test. ssGSEA scores for gene programs were z-transformed across samples and analyzed using the Wilcoxon rank-sum test. When comparing paired samples from the patients (eg pre-trastuzumab vs post-trastuzumab), paired t test was employed. Spatial autocorrelation of *ERBB2* expression within each HER2-positive sample was assessed using Moran's *I* test, implemented via the R package spdep (v.1.2-8). Differential gene expression was performed using LLM for DSP data. Multiple comparisons were adjusted using the false discovery rate (FDR) method.

**Data Availability**

Raw sequencing data of DSP profiles have been deposited at the European

Genome-phenome Archive (EGA), which is hosted by the EBI and the CRG, under accession number EGAS50000000636. Further information about EGA can be found at https://ega-archive.org and "The European Genome-phenome Archive of human data consented for biomedical research". Additional datasets employed in the study are detailed in their respective sections of the Methods.
